# An in-depth analysis of phosphorylated tau in amyotrophic lateral sclerosis post-mortem motor cortex and cerebrospinal fluid

**DOI:** 10.1101/2020.12.30.20248944

**Authors:** Tiziana Petrozziello, Ana C. Amaral, Simon Dujardin, Sali M.K. Farhan, James Chan, Bianca A. Trombetta, Pia Kivisäkk, Alexandra N. Mills, Evan A. Bordt, Spencer E. Kim, Patrick M. Dooley, Anubrata Ghosal, Teresa Gomez-Isla, Bradley T. Hyman, Steven E. Arnold, Tara Spires-Jones, Merit E. Cudkowicz, James D. Berry, Ghazaleh Sadri-Vakili

## Abstract

Although the molecular mechanisms underlying amyotrophic lateral sclerosis (ALS) are not yet fully understood, recent studies have described alterations in tau protein in both sporadic and familial ALS. However, it is unclear whether alterations in tau contribute to ALS pathogenesis. Here, we leveraged the ALS Knowledge Portal and Project MinE data sets and identified specific genetic variants clustering within the microtubule-binding domain of *MAPT*, which were unique to ALS cases. Furthermore, our analysis in a large post-mortem cohort of ALS and control motor cortex demonstrates that although there was no significant difference in the presence of phosphorylated tau (pTau) neuropil threads and neurofibrillary tangles between the two groups, pTau-S396 and pTau-S404 mis-localized to the nucleus and synapses in ALS. This was specific to the C-terminus phosphorylation sites as there was a significant decrease in pTau-T181 in ALS synaptoneurosomes compared to controls. Lastly, while there was no change in total tau or pTau-T181 in ALS CSF, there was a decrease in pTau-T181:tau ratio in ALS CSF, as previously reported. Importantly, CSF tau levels were increased in ALS patients diagnosed with bulbar onset ALS, while pTau-T181:tau ratio was decreased in ALS patients diagnosed with both bulbar and limb onset. Additionally, there was an inverse correlation between tau levels in the CSF and the revised ALS functional rating scale (ALSFRS-R) as well as a correlation between pTau-T181:tau ratio and ALSFRS-R. While there were no longitudinal alterations in tau, pTau-T181 and pTau-T181:tau ratio, there was an increase in the rate of ALSFRS-R decline per month associated with increases in tau levels. This decline was also inversely correlated with increases in pTau-T181 in relation to tau levels. Taken together, our findings demonstrate that, like Alzheimer’s disease, hyperphosphorylated tau is mis-localized in ALS and that decreases in CSF pTau-T181 may serve as a biomarker in ALS.

## Introduction

Amyotrophic lateral sclerosis (ALS) is a fatal neurodegenerative disease that primarily affect both cortical (upper) and spinal (lower) motor neurons [1-2]. Several genes have been implicated in ALS pathogenesis [3-4], however mutations in these genes account for a minority of cases, and the etiology of the disease remains to be elucidated. Therefore, understanding the exact molecular mechanisms leading to motor neuron loss is crucial for the development of both new therapeutic approaches and the discovery of novel and useful biomarkers of disease.

Recent studies have begun to link alteration in tau phosphorylation to ALS pathogenesis with tau pathology reported in both sporadic and familial cases [5-6]. Tau protein is a member of the microtubule-associated protein (MAP) family and plays a critical role in stabilizing microtubules, the major component of the eukaryotic cytoskeleton involved in cell processes, including cell division, mobility, and the intracellular organization and trafficking of organelles [7]. Tau hyperphosphorylation, accumulation, and mutations have been linked to a group of progressive neurodegenerative diseases collectively known as tauopathies [8-11], in which hyperphosphorylation of key epitopes on tau promote its disassembly from microtubules, aggregation, and subcellular mis-localization, leading to the formation of inclusions and neurofibrillary tangles (NFTs) in both neurons and glia [9, 12-13].

Although the exact molecular mechanisms underlying tau toxicity are not yet fully understood, the main consequence of the accumulation of toxic tau is the disruption of neuronal transport [14-17]. This impairment is an early pathogenic event in neurodegeneration and tau-mediated alterations in neuronal trafficking have been described in several neurodegenerative diseases [14-17]. Furthermore, deficits in neuronal transport were shown to disrupt several cellular functions, including but not limited to alterations in both trafficking and function of mitochondria, synapse loss, excitotoxicity, and cell death [14-15, 17-18].

In ALS there is a significant increase in total tau as well as cytoplasmic inclusions of hyperphosphorylated tau (T175, T217, S208/210, S212, S396 and S404) in post-mortem motor cortex (mCTX) and spinal cord of ALS patients [19-21]. Moreover, alterations in total tau and pTau:tau ratio have been reported in the cerebrospinal fluid (CSF) of ALS patients [22-25]. Importantly, tau-induced alterations in cellular processes such as excitotoxicity, mitochondrial dysfunction, synapse loss, and impairments of nucleocytoplasmic transport, are also pathogenic features of ALS [1, 26-28], suggesting that alterations in tau could underlie these molecular events in ALS.

Here, we sought to determine whether there were novel genetic variants in *MAPT*, the gene encoding tau, in ALS patients. Furthermore, we used a large cohort of ALS post-mortem motor cortex (mCTX) to investigate alterations in tau and its possible molecular implications in ALS. Lastly, we measured CSF tau, pTau-T181, and pTau-T181:tau ratio in ALS patients and healthy controls given the previous contradictory results from previous biomarker studies in ALS.

## Material and Methods

All methods were carried out in accordance with the guidelines and regulations of Massachusetts General Hospital and approved by the Massachusetts General Hospital licensing and ethics committees.

### Identifying MAPT variants in ALS patients

The genetic approach taken herein was previously described by Petrozziello et al. [29]. Briefly, genetic data were obtained from the two largest repositories on ALS patients, ALS Knowledge Portal (ALSKP) [30] and Project MinE data browser [31]. We included any variants annotated as missense, non-synonymous, and splice altering variants within ALS cases (ALSKP, n=3,864 cases and n=7,839 controls; Project MinE, n=4,366 cases and n=1,832 controls). We used the genome Aggregation Database (gnomAD, 125,748 exomes and 15,708 genomes, total n=141,456) [32] to determine the global population frequency of each variant. For all the variants observed, we included the annotation outputs derived from CADD and MPC, which help guide us during variant interpretation and prioritization. All variants are displayed in Table 1. Variant positions are based on reference genome assembly GRCh37/hg19. We also surveyed ClinVar, a repository of genetic variants reported in patients with disease for any *MAPT* pathogenic or likely pathogenic variants. Finally, we used *MAPT* isoform NM_016835 to report the genetic variants in accordance with gnomAD and ClinVar.

**Table 1.** MAPT variants in ALS.

| Source | Variant | Protein change | Consequence | ALS cases allele count | Controls allele count | gnomAD AF | gnomAD AF (non-neuro) | MPC | CADD | ClinVar | ALS type |
| --- | --- | --- | --- | --- | --- | --- | --- | --- | --- | --- | --- |
| ALSKP | 17:44039728G>A | p.Glu9Lys | missense | 1 | 1 | 1.59E-05 | 1.92E-05 | 0.18 | 14.21 | NR | ALS rare |
| ALSKP | 17:44039730A>C | p.Glu9Asp | missense | 1 | 0 | 0 | 0 | 0.13 | 0.011 | NR | ALS unique |
| ALSKP | 17:44039796G>C | p.Met31Ile | missense | 1 | 0 | 3.99E-06 | 0 | 0.17 | 16.9 | NR | ALS unique |
| Project MinE | 17:44049250G>C | p.Glu53Asp | missense | 1 | 0 | 3.98E-06 | 0.00E+00 | 0.14 | 13.39 | NR | ALS unique |
| ALSKP | 17:44049250G>C | p.Glu53Asp | missense | 1 | 0 | 3.98E-06 | 0 | 0.14 | 13.39 | NR | ALS unique |
| Project MinE | 17:44051801G>A | p.Ala91Thr | missense | 2 | 1 | 2.19E-05 | 1.99E-05 | 0.36 | 24.7 | VUS | ALS rare |
| Project MinE | 17:44051807C>T | p.Pro93Ser | missense | 1 | 0 | 0.00E+00 | 0.00E+00 | 0.16 | 15.73 | NP | ALS unique |
| Project MinE | 17:44055752G>A | p.Gly107Ser | missense | 1 | 0 | 1.19E-05 | 9.61E-06 | 0.65 | 26 | NR | ALS rare |
| Project MinE | 17:44055753G>T | p.Gly107Val | missense | 2 | 1 | 1.19E-05 | 1.44E-05 | 0.73 | 26.6 | VUS | ALS rare |
| Project MinE | 17:44060867C>T | p.Pro233Ser | missense | 1 | 0 | 7.23E-06 | 8.92E-06 | 0.60 | 17.71 | NR | ALS rare |
| ALSKP | 17:44061026G>A | p.Gly286Arg | missense | 1 | 0 | 1.77E-05 | 1.44E-05 | 0.18 | 0.215 | NR | ALS rare |
| ALSKP | 17:44061109G>T | p.Lys313Asn | missense | 1 | 1 | 1.59E-05 | 1.93E-05 | 0.57 | 23.5 | Likely benign | ALS rare |
| ALSKP | 17:44061117C>T | p.Ala316Val | missense | 1 | 0 | 3.99E-06 | 4.82E-06 | 0.15 | 5.652 | NR | ALS rare |
| Project MinE | 17:44061117C>T | p.Ala316Val | missense | 1 | 0 | 3.99E-06 | 4.82E-06 | 0.15 | 5.652 | NR | ALS rare |
| Project MinE | 17:44061182C>T | p.Arg338Trp | missense | 1 | 0 | 0.00E+00 | 0.00E+00 | 0.16 | 23 | NR | ALS unique |
| Project MinE | 17:44061185G>A | p.Gly339Ser | missense | 2 | 0 | 3.19E-05 | 4.71E-05 | 0.17 | 8.896 | NR | ALS rare |
| Project MinE | 17:44061261G>A | p.Arg364Gln | missense | 1 | 0 | 1.79E-05 | 2.18E-05 | 0.18 | 11.85 | NR | ALS rare |
| Project MinE | 17:44061272G>C | p.Val368Leu | missense | 1 | 0 | 0.00E+00 | 0.00E+00 | 0.15 | 6.996 | NR | ALS unique |
| Project MinE | 17:44064409G>T | p.Arg377Leu | missense | 1 | 0 | 3.98E-06 | 4.81E-06 | 0.64 | 24.8 | NR | ALS rare |
| ALSKP | 17:44064432G>A | p.Gly385Arg | missense | 1 | 1 | 2.12E-05 | 2.18E-05 | 0.23 | 22.5 | NR | ALS rare |
| ALSKP | 17:44067307A>G | p.Thr416Ala | missense | 1 | 1 | 2.48E-05 | 2.62E-05 | 0.32 | 19.68 | NR | ALS rare |
| ALSKP | 17:44067335ACCC>- | p.Pro426del | inframe deletion | 1 | 0 | 0 | 0 | NA | NA | NR | ALS unique |
| ALSKP | 17:44067353T>C | p.Val431Ala | missense | 1 | 0 | 0 | 0 | 0.54 | 22.8 | NR | ALS unique |
| Project MinE | 17:44067353T>C | p.Val431Ala | missense | 1 | 0 | 0.00E+00 | 0.00E+00 | 0.54 | 22.8 | NR | ALS unique |
| ALSKP | 17:44067447G>C | c.1380+6G>C | splice region | 1 | 0 | 1.19E-05 | 4.81E-06 | NA | 16.27 | NR | ALS rare |
| ALSKP | 17:44068857C>T | p.Pro471Leu | missense | 1 | 0 | 3.98E-06 | 0 | 0.65 | 26 | NR | ALS unique |
| Project MinE | 17:44073789G>A | p.Arg529His | missense | 1 | 0 | 4.13E-06 | 0.00E+00 | 2.26 | 28.2 | NR | ALS unique |
| Project MinE | 17:44073809G>A | p.Gly536Ser | missense | 1 | 0 | 2.93E-05 | 3.18E-05 | 2.07 | 29.3 | NR | ALS rare |
| Project MinE | 17:44074004C>G | p.Leu583Val | missense | 2 | 0 | 0.00E+00 | 0.00E+00 | 1.94 | 24.9 | Pathogenic | ALS unique |
| Project MinE | 17:44087690TAAG>T | p.Lys616del | inframe deletion | 1 | 0 | 2.62E-05 | 3.13E-05 | NA | NA | Conflicting | ALS rare |
| ALSKP | 17:44091643A>T | p.Lys652Met | missense | 2 | 0 | 0 | 0 | 2.32 | 27.3 | Pathogenic | ALS unique |
| ALSKP | 17:44096073G>A | p.Val698Ile | missense | 1 | 0 | 2.83E-05 | 3.05E-05 | 0.76 | 21.3 | Conflicting | ALS rare |
| ALSKP | 17:44101439A>C | p.Asn745His | missense | 1 | 1 | 1.19E-05 | 4.81E-06 | 2.10 | 24.4 | NR | ALS rare |
| ALSKP | 17:44101466A>G | p.Met754Val | missense | 1 | 0 | 3.98E-06 | 0 | 1.77 | 22.9 | NR | ALS unique |
| ALSKP | 17:44101481C>A | p.Gln759Lys | missense | 1 | 1 | 2.79E-05 | 2.89E-05 | 1.86 | 27 | NP | ALS rare |
| ALSKP | 17:44101487G>A | p.Ala761Thr | missense | 2 | 2 | 2.48E-05 | 2.18E-05 | 1.69 | 24.2 | NR | ALS rare |

Abbreviations are as follows: ALSKP, ALS Knowledge Portal (ALSKP.org); AF, allele frequency; NR, not reported; VUS, variant of uncertain significance; conflicting, conflicting interpretations; NP, not provided. In this context, ALS type refers to whether the variant has only been observed in ALS cases (ALS unique) or has been observed in gnomAD, but at a very low allele frequency (ALS rare).

### Human tissue samples

Post-mortem motor cortices from control and ALS patient brains were provided by the Massachusetts Alzheimer’s Disease Research Center (ADRC) with approval from the Massachusetts General Hospital Institutional Review Board (IRB). In total we assessed 50 ALS and 25 non-neurological control motor cortices, and entorhinal cortex (EC) from two positive controls with Alzheimer’s disease (AD) (provided by the MGH ADRC). The mean age was 73.0 years (SD = 14) for control subjects and 63.28 years (SD = 10) for ALS subjects. For the control group there was 48% male, while there was 58% male in the ALS group. Twenty-two ALS patients were diagnosed with limb onset disease, while 15 patients were diagnosed with bulbar onset. Three were diagnosed with ALS/frontotemporal dementia (ALS/FTD). Five of the ALS patients were positive for *C9ORF72* repeat expansion, while a single case was positive for an *SOD1* mutation. Genetic status of all other ALS subjects was unknown. Thirty of the ALS cases showed TDP-43 proteinopathy and three were diagnosed with ALS/FTD. Post-mortem interval (PMI) range was 9-86 h for control subjects and 4-77 h for ALS subjects.

### Human cerebrospinal fluid (*CSF*) *samples*

After obtaining written informed consent, cerebrospinal fluid samples were obtained from participants with ALS (n = 40) and controls (n = 10) at the Healey Center for ALS at Mass General. Longitudinal CSF samples and accompanying clinical information from participants with ALS were collected between 2011 and 2016 as a part of a prospective, multicenter observational study. Control samples were obtained in a concurrently enrolling single-center study at MGH using identical techniques to obtain, process, store, and share biofluid samples. In each study, participants were enrolled, detailed clinical information and CSF samples were obtained at baseline and, for longitudinally collected samples, at follow-up visits, approximately every 4 months. Cerebrospinal fluid was centrifuged, aliquoted, and frozen at −80°C. Processing was initiated within 15 minutes of collection. Clinical data collected from participants with ALS included timing and location of disease onset and progression, including the revised ALS Functional Rating Scale (ALSFRS-R) and slow vital capacity (SVC). Raters for the ALSFRS-R and VC were trained by the Northeast ALS Consortium (NEALS) Outcomes Training Center at the Barrow Neurological Institute. CSF sample information is provided in Table 2.

**Table 2.** Demographics of ALS CSF samples.

|  | Age at sample collection<br>[mean (SD)] | Disease duration<br>[months, mean, (SD)] | ALSFRS-R total score<br>[mean (SD)] | Pre-baseline<br>ALSFRS-R slope<br>[mean (SD)] |
| --- | --- | --- | --- | --- |
| Visit 1 | n=40, 52.41 (12.32) | n=33, 24.41 (19.05) | n=40, 37.92 (6.41) | n=33, 0.57 (0.50) |
| Visit 2 | n=26, 56.52 (11.18) | n=19, 33.33 (22.54) | n=26, 36.00 (6.82) | n=19, 0.37 (0.24) |
| Visit 3 | n=22, 55.82 (10.81) | n=17, 38.60 (24.35) | n=22, 34.27 (6.74) | n=17, 0.34 (0.23) |
| Visit 4 | n=16, 58.25 (8.91) | n=12, 48.45 (27.91) | n=16, 32.94 (8.19) | n=12, 0.30 (0.22) |
| Visit 5 | n=12, 55.59 (11.32) | n=10, 46.80 (15.01) | n=12, 30.75 (5.40) | n=10, 0.31 (0.17) |
| Visit 6 | n=10, 54.87 (11.11) | n=8, 55.30 (16.52) | n=10, 27.70 (7.13) | n=8, 0.27 (0.15) |
| Visit 7 | n=5, 62.08 (11.47) | n=4, 58.95 (19.43) | n=5, 25.40 (8.68) | n=4, 0.30 (0.19) |

### Immunohistochemistry and Image Analysis

Seven-μm-thick paraffin-embedded brain sections from the motor cortex (Broadmann area 4) and the entorhinal cortex were immunostained for pTau-S396 (1:100; Abcam, MA) and pTau-S396/S404 (PHF1; 1:250; Peter Davies) using a Bond Rx autostainer (Leica Biosystems, IL), according to the manufacturer’s instructions. Briefly, slides were batch processed with the following settings: Bake and Dewax, IHC protocol F 60 minutes, HIER 20 minutes with ER1. Slides were then transferred into water and dehydrated by 1-minute incubations into baths of 70% ethanol, 95% ethanol, 100% ethanol, and xylene. Slides were then cover slipped using Permount Mounting Medium (Fisher Scientific) and left to dry overnight. Slides were scanned using an Olympus VS120 virtual slide microscope at a magnification of 20X. Scanned slide images were analyzed in Olympus cellSens and OlyVIA analysis softwares.

### Nuclear/cytosolic fractionation

Nuclear and cytosolic fractions were prepared from control and ALS post-mortem mCTX using the CelLytic NuCLEAR Extraction kit (#NXTRACT, Sigma-Aldrich, MO), according to the manufacturer’s instruction and as previously described [33]. Briefly, 100 mg tissue was homogenized in hypotonic lysis buffer supplemented with 0.1 M dithiothreitol (DTT) and protease inhibitors and centrifuged at 10,000 g for 20 min. The supernatants were collected and saved as the cytosolic fraction. The remaining pellets were resuspended in extraction buffer containing 0.1 M DTT and protease inhibitors, and gently agitated for 30 min at 4°C. Next, samples were centrifuged at 20,000 g for 5 min, and supernatants were collected and saved as the nuclear fraction. Protein concentration was determined by Bradford assay.

### Synaptoneurosome (SNs) fractionation

SNs fractionation was performed as previously reported [34] with few modifications. Human post-mortem mCTX (200 to 300 mg) were homogenized in 500 μL ice-cold Buffer A composed of 25 mM Hepes pH 7.5, 120 mM NaCl, 5 mM KCl, 1 mM MgCl_2_, 2 mM CaCl_2_, 2 mM dithiothreitol (DTT), 1 mM NaF supplemented with phosphate and protease inhibitors cocktail. The homogenates were then passed through two layers of 80 μm nylon filters (Millipore, MA) to remove tissue debris. Seventy μL aliquot was saved, mixed with 70 μL H_2_O and 23 μL 10% sodium dodecyl sulphate (SDS), passed through a 27-gauge needle to shear DNA, boiled for 5 min, and centrifuged at 15,000 g for 15 min to prepare the total extract. The remainder of the homogenates was passed through a 5 μm Supor membrane Filter (Pall Corp, Port Washington, NY) to remove organelles and nuclei, centrifuged at 1000 g for 5 min and both pellet and supernatant were saved. The supernatants were collected in small crystal centrifuge tubes and centrifuged at 100,000 g for 45 min at 4^°^C to obtain the cytosolic fractions. The pellets were resuspended in Buffer B composed of 50 mM Tris pH 7.5, 1.5% SDS and 2 mM DTT, boiled for 5 min, and centrifuged at 15,000 g for 15 min to obtain SNs. Bradford assay was used to determine protein concentration in total extract, cytosolic fraction and SNs.

### Western blotting

Western blots were performed using previously described protocols [33, 35]. Briefly, 50 μg of proteins was resuspended in sample buffer and fractionated on a 4-12% bis-tris gel for 90 min at 120V. Proteins were then transferred to a PVDF membrane in an iBlot Dry Blotting System (Invitrogen, Thermo Fisher, MA), and the membrane was blocked with 5% bovine serum albumin (BSA) in tris-buffered saline with Tween 20 (TBST) before immunodetection with the following primary antibodies: pTau-S396 (1:500; Abcam, MA), pTau-S404 (1:500; Abcam, MA), pTau-T181 (1:500; Abcam, MA), tau (1:1000, DAKO, Denmark), histone H3 (1:500; Millipore, MA), PSD-95 (1:1000, Cell Signaling, MA), Β-Actin (1:1000; Cell Signaling, MA), and GADPH (1:1000; Millipore Sigma, Termecula, CA), overnight at 4°C. Primary antibody incubation was followed by 4 washes in TBST before incubation with the secondary antibody for 1 h (HRP-conjugated goat anti-rabbit IgG, and HRP-conjugated goat anti-mouse IgG; Jackson ImmunoResearch Laboratories, West Grove, PA). After 4 washes in TBST, proteins were visualized using the ECL detection system (Thermo Fisher Scientific, MA).

### Quanterix Simoa Assays

CSF tau and pTau-T181 concentrations were measured using the Simoa Tau and pTau181 Advantage Kits on a fully automated Simoa HD-X Analyzer using manufacturer’s recommendations (Quanterix Corporation, Billerica, MA). CSF samples were centrifuged at 3,000 g for 10 minutes, diluted 1:4 (pTau-T181) or 1:10 (tau) in sample buffer, and run in duplicate. The coefficient of variance (CV) was 0-8% (mean±SD = 2.2±1.9%) for pTau-T181 and 0-26% (mean±SD = 5.4±5.2%) for tau. One sample with high CV (77%) was excluded from further analysis. The lower limit of quantification (LLOQ) was 4.8 pg/ml for pTau-T181 and 0.75 pg/ml for tau.

### Statistics

Normal distribution of data were not assumed regardless of sample size or variance. The data are presented as individual value plots with the central line representing the median and the whiskers represented the interquartile range or box plots with the central line representing the median, the edges representing the interquartile ranges, and the whiskers representing the minimum and maximum values. For CSF analysis, pre-baseline ALSFRS-R slope was calculated as the [(48 – ALSFRS-R score) / (months since disease onset)] where ALSFRS-R score is the first ALSFRS-R total score recorded. Trajectory of ALSFRS-R total score was estimated using a mixed effects model with a fixed effect for time and a random intercept and slope for each subject with an unstructured covariance. A covariate for each CSF measure was added separately to the ALSFRS-R total score trajectory model to describe any effect on change in ALSFRS-R total score. Changes in CSF measures were compared to changes in ALSFRS-R total score by taking the first value for each subject subtracted from the last available visit with complete data for the given comparison. Comparisons between groups were performed using a non-parametric Mann-Whitney U test, a one-way ANOVA followed by Tukey’s post-hoc test, a two-way ANOVA followed by Tukey’s post-hoc test, and a Fisher’s exact test followed by Bonferroni correction for multiple comparison for semi-quantitative scores. Correlations of CSF measures with clinical measures were performed as non-parametric Spearman correlations. Comparisons for clinical measures were not corrected for multiple comparisons. All tests were two sided with a significance level of 0.05, and exact p values are reported. Analyses were performed using GraphPad Prism and R, a language and environment for statistical computing (https://www.R-project.org/).

### Study approval

The study was approved by the Partners Healthcare Institutional Review Board (IRB). Written informed consent was obtained from all participants prior to study enrollment. Post-mortem consent was obtained from the appropriate representative (next of kin or health care proxy) prior to autopsy.

## Results

### MAPT genetic variants in ALS patients

Though pathogenic *MAPT* variants have been established as a cause of frontotemporal dementia related neurodegeneration, herein, via our genomics approach, we also observed multiple *MAPT* variants in ALS cases. Specifically, we observed a total of 36 heterozygous variants in the *MAPT* gene in a total of 42 ALS cases (Table 1) and no homozygous variants aggregated in ALSKP and Project MinE, the two largest ALS repositories of genomic data (ALSKP, n = 3,864 cases and n = 7,839 controls; Project MinE, n = 4,366 cases and n = 1,832 controls). Among the 36 genetic variants, 33 were missense variants (Figure 1A). Next, we evaluated the population frequency of each *MAPT* variant using large datasets aggregating exomes and genomes from non-ALS individuals such as genome Aggregation Database (gnomAD). Among the 36 variants, 15 variants were unique to ALS cases and absent in gnomAD and were accordingly classified as ‘ALS unique variants’. Similarly, 21 variants were classified as ‘ALS rare variants’ as they were observed in gnomAD at a very low frequency (MAF < 4.71E-05) (Figure 1B). Following variant annotation with CADD and MPC, we divided the missense variants into 5 distinct categories (M1-M5) based on the predicted severity of their impact on protein function with M1 (MPC > 2 and CADD > 25) being the most probable to induce a pathogenic effect and M5 being the least probable category (MPC < 1.5 and CADD < 20). The variants in categories M1 and M2, which are within the upper limit of predicted pathogenicity, cluster near the C-terminus of the protein transcript within or around the microtubule-binding domain (Figure 1B). The addition of known pathogenic or likely pathogenic *MAPT* variants reported in ClinVar in patients with neurodegenerative diseases, specifically frontotemporal dementia and other clinically overlapping diseases such as Parkinson’s disease, late-onset Parkinson-dementia syndrome, Pick’s disease, and progressive supranuclear ophthalmoplegia, further supports the predicted pathogenicity of M1 and M2 variants as these ClinVar variants also cluster at the C-terminus within or neighboring the tubulin-binding domain (Figure 1C). Of four ALS variants that are harbored within the tubulin-binding domain namely, p.Leu583Val, p.Lys616del, p.Lys652Met, and p.Val698Ile, two variants were previously reported in ClinVar. One variant, p.Leu583Val, was also reported in a patient with FTD, and classified as a pathogenic variant, the most severe consequence assigned to a variant. The brain of the patient carrying the variant, which can also be written as p.Leu266Val (depending on the isoform type), showed Pick body-like inclusions and unique tau-positive, argyrophilic astrocytes with stout filaments and naked, round, or irregular argyrophilic inclusions with deposits of both three-repeat and four-repeat tau [36]. The variant was observed in two unrelated ALS patients in the Project MinE browser, which does not include phenotypic information. Therefore, it remains unclear whether these ALS patients may develop additional symptoms consistent with FTD. Furthermore, another variant, p.Lys652Met, which is unique to ALS cases and was also observed in two unrelated ALS patients, has been previously reported in ClinVar in two pedigrees displaying FTD and ALS [37]. The patients were described to have parkinsonism and pyramidalism, with half of them having amyotrophy. In addition, neuropathologically, an extensive deposition of abnormal tau protein in a mixed pattern (neuronal, glial) was observed. For the other variants within and outside of the tubulin-binding domain, despite their rarity and even several being unique to ALS cases, their pathogenicity and contribution to neurodegeneration remains elusive, and their effect would need to be validated by observing them in the disease state or modelled *in vitro*.

**Figure 1.**
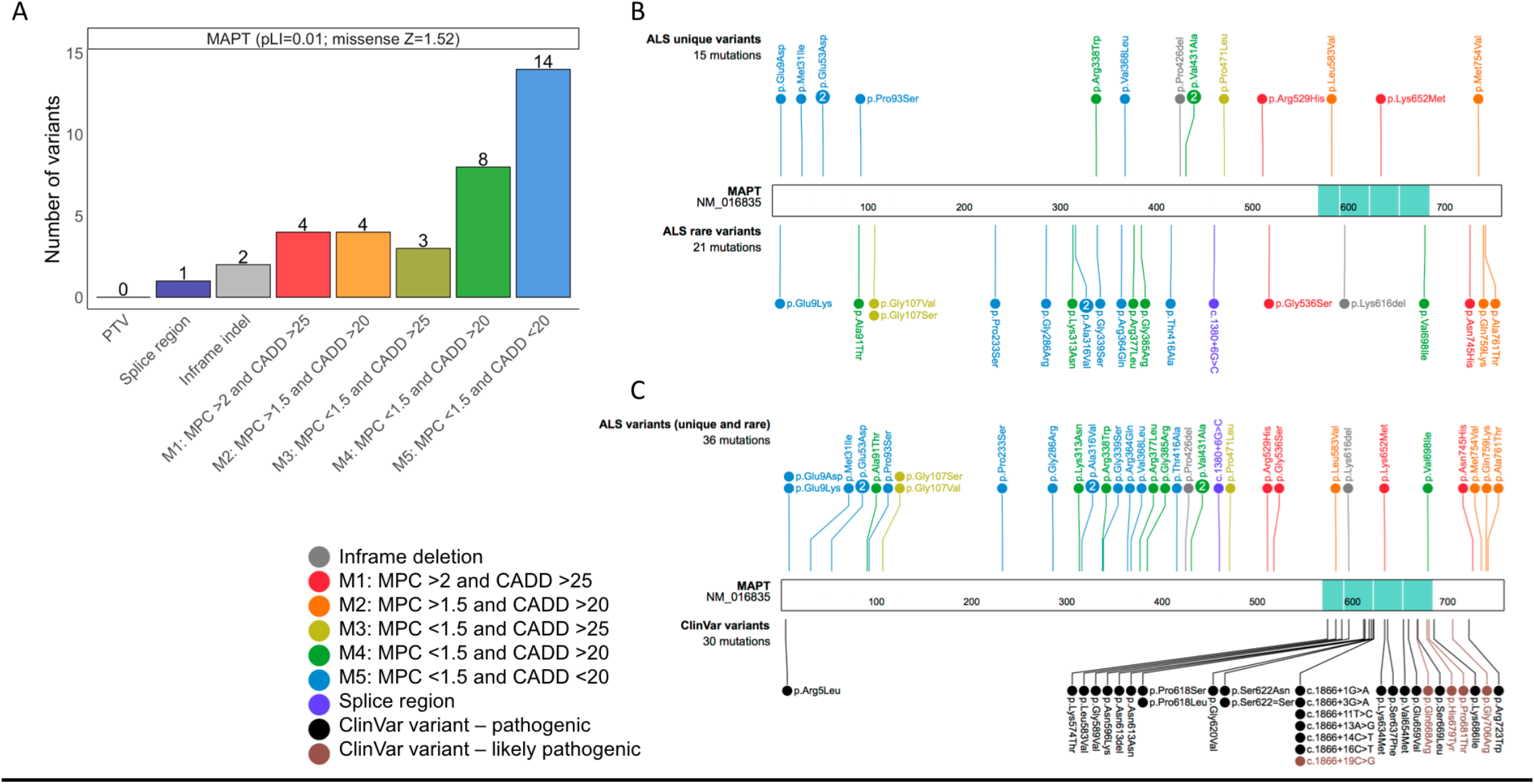
Protein schematic of *MAPT* variants observed in ALS cases. **(A)** Distribution of variant type in *MAPT* in ALS cases. Variant types are displayed on the X-axis with their respective counts on the Y-axis for the *MAPT* gene. The exact number of variants observed is noted on top of each bar. The probability of loss of function (pLI) and the missense constraint Z scores for *MAPT* are shown adjacent to the gene label. PTV, protein truncating variant; indel, insertion/deletion; M in M1-M5, missense. Missense variants were divided into 5 classes depending on their MPC and CADD scores. **(B)** The variants displayed on top are variants identified only in ALS cases and are classified as ‘ALS unique variants’. The variants displayed on the bottom were observed in ALS cases and also at a very low allele frequency in gnomAD, therefore these were classified as ‘ALS rare variants’. The colors represent the type of non-synonymous changes observed, and the number next to the variants depicts the number of individuals observed to carry the corresponding variant. M1-M5: missense variant with different MPC and CADD thresholds. The tubulin-binding domain is shown in turquoise. The numbers within the protein sequence depicts the amino acid position. **(C)** Schematic representation of all ALS variants (unique and rare) on top and ClinVar pathogenic (black) and likely pathogenic (brown) variants on the bottom.

### Tau and pTau levels are not changed in ALS post-mortem motor cortex (mCTX)

To determine whether tau or phosphorylated tau (pTau) levels are altered in ALS, we used a large cohort of post-mortem samples and assessed the levels of tau, pTau-S396, pTau-S404, and pTau-T181 in post-mortem ALS (n = 36) and control (n = 14) mCTX by western blots. The results demonstrate that there were no significant alterations in tau (Mann Whitney U = 171, p = 0.1588), pTau-S396 (Mann Whitney U = 181, p = 0.2898), pTau-S404 (Mann Whitney U = 213, p = 0.7484), or pTau-T181 (Mann Whitney U = 204.5, p = 0.7027) levels in ALS mCTX compared to controls (Figure 2A-E).

**Figure 2.**
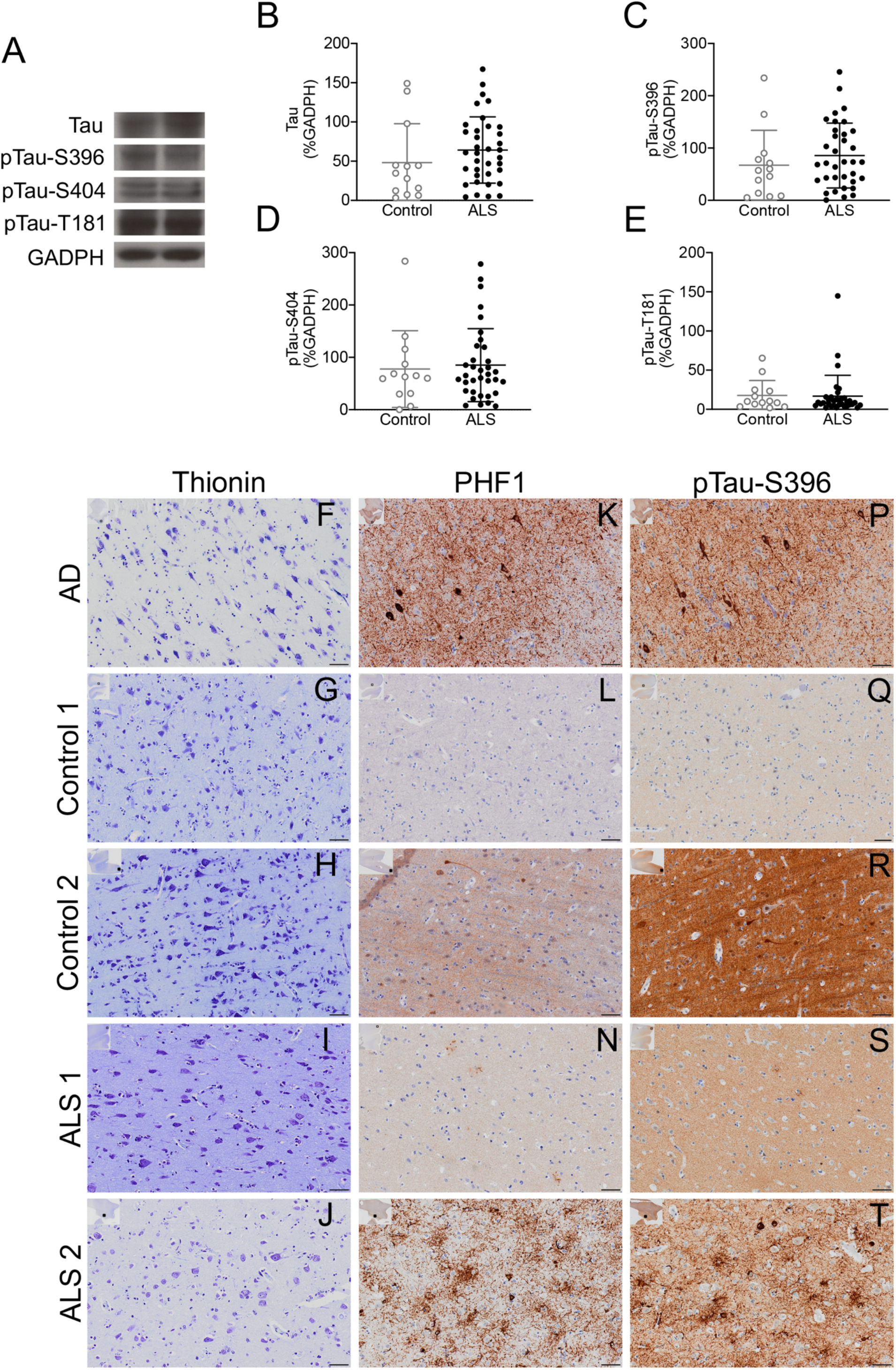
pTau levels are not altered in ALS post-mortem mCTX. **(A)** Representative western blot images of tau, pTau-S396, pTau-S404, and pTau-T181 in ALS and control mCTX. **(B)** There was no change in tau levels in ALS mCTX compared to controls (Mann Whitney U = 171, p = 0.1588). **(C)** pTau-S396 levels were not altered in ALS mCTX compared to controls (Mann Whitney U = 181, p = 0.2898). **(D)** There was no alteration in pTau-S404 levels in ALS compared to control mCTX (Mann Whitney U = 213, p = 0.7484). **(E)** There was no change in pTau-T181 levels in ALS mCTX compared to controls (Mann Whitney U = 204.5, p = 0.7027). (**F-J**) Thionin immunostaining in AD, control, and ALS post-mortem mCTX. (**K**) PHF1 immunostaining in AD EC demonstrating intense neuropil threads and NFTs. Representative images of weak (+) PHF1 immunostaining in control (**L**) and ALS (**N**) mCTX. Representative images of intense (+++) PHF1 immunostaining in control (**M**) and ALS (**O**) mCTX. (**P**) pTau-S396 immunostaining in AD EC demonstrating intense neuropil threads and NFTs. Representative images of weak (+) pTau-S396 immunostaining in control (**Q**) and ALS (**S**) mCTX. Representative images of intense (+++) pTau-S396 immunostaining in control (**R**) and ALS (**T**) mCTX. The graphs demonstrate the individual integrated density values (IDV) for tau, pTau-S396, pTau-S404, and pTau-T181 as % of GADPH IDV from western blot experiments performed in mCTX from ALS (n = 36) and non-neurological control (n = 14), with the central line representing the median, and the edges representing the interquartile range. The images represent thionin, PHF1, and pTau-S396 immunostaining from IHC experiments performed in AD EC (n = 2), control (n = 8) and ALS mCTX (n = 14 for PHF1 staining, and n = 18 for pTau-S396 staining). Scale bar: 50 μM

To confirm the findings from the western blot analysis, we assessed pTau levels in ALS (n = 14) and non-neurological control (n = 8) mCTX by immunohistochemistry (IHC) using PHF-1 antibody, able to recognize pTau at both S396 and S404 (kindly provided by Dr. Peter Davies). Nissl body marker thionin was used to visualize cells in the adjacent sections from the same post-mortem ALS and control mCTX samples (Figure 2F-J) and two AD post-mortem EC were included as positive controls. PHF-1 immunosignal was detected in both control, ALS and AD samples. As expected, extensive neuropil threads and NFTs were observed in AD brains (Figure 2K). Both control (Figure 2L and M) and ALS mCTX (Figure 2N and O) demonstrated heterogeneity of PHF1 immunosignal, therefore, we categorized the intensity of the signal as outlined in Table 3. Intense (+++) PHF1 immunostaining was detected in 7.1% of ALS mCTX, while moderate (++) PHF1 staining was observed in 14.3% of ALS and 37.5% of control mCTX, and weak (+) staining in 35.7% of ALS and 25% of control cases. Similarly, extensive (+++) neuropil threads were detected in 7.1% of ALS cases, moderate (++) neuropil threads in 21.4% of ALS and 25% of control cases, and weak (+) neuropil threads in 14.3% of ALS and 37.5% of control cases. Moreover, when we assessed the presence of NFTs using PHF1, 7.1% of ALS cases demonstrated a high number (+++) of NFTs in 14.3% of ALS and 12.5% of control cases. While moderate numbers of NFTs (++) were detected in 28.6% of ALS cases, 37.5% of control cases exhibited few and sparse NFTs (+). Lastly, PHF1 staining was absent (-) in 42.9% of ALS and 37.5% of controls, neuropil threads were absent (-) in 57.1% of ALS and 37.5% of control cases, and no NFTs (-) were detected in 50% of ALS and 50% of control cases (Table 3). Furthermore, the statistical analysis of the semi-quantitative immunosignal scores revealed no significant difference in PHF1 overall staining, neuropil threads, and NFTs between ALS and controls (Fisher’s test, p = 0.7564, p = 0.7523, and p = 1, respectively).

**Table 3.**
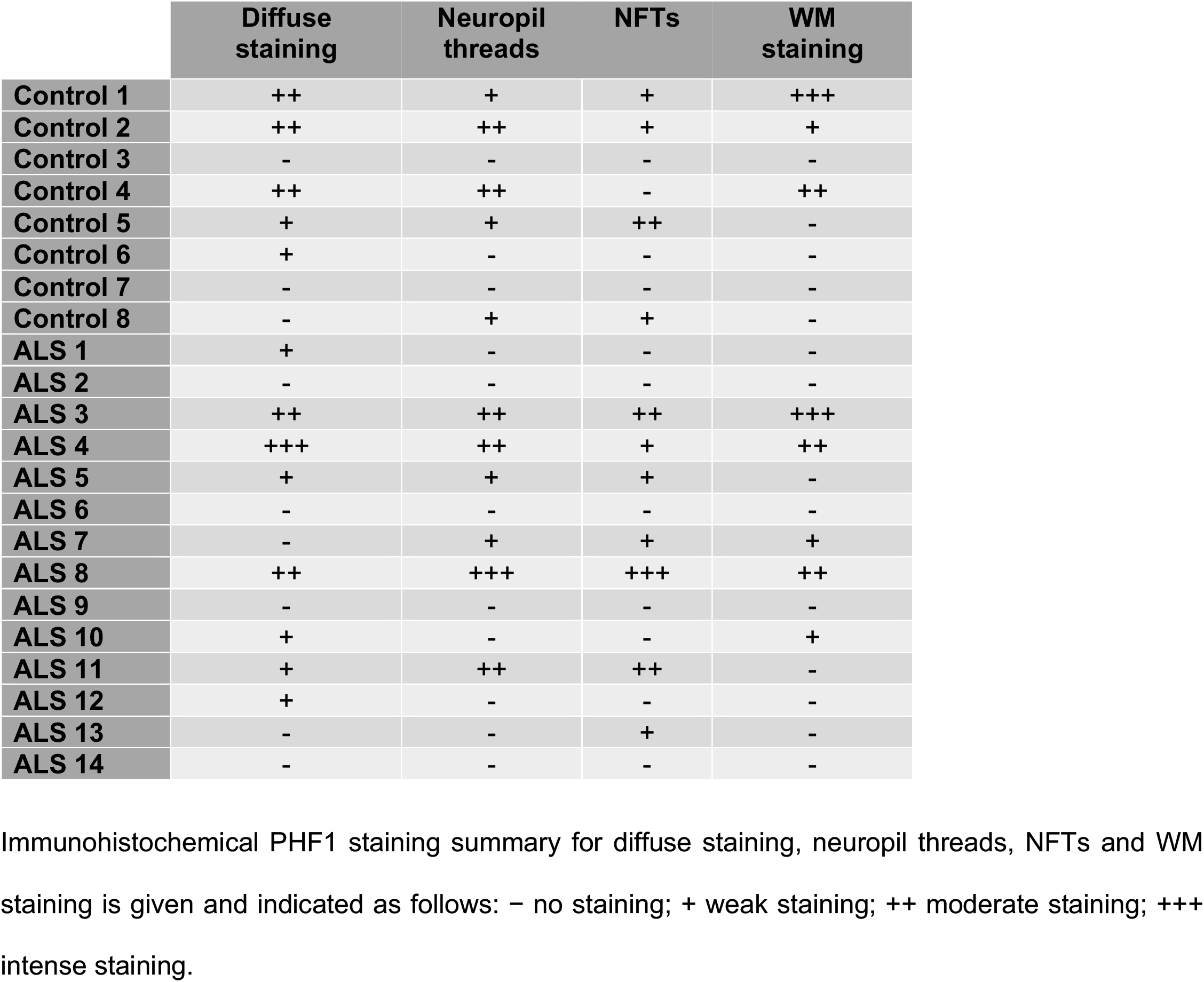
PHF1 immunostaining in control and ALS post-mortem mCTX.

We further confirmed our findings by assessing pTau-S396 levels alone in a total of 18 ALS samples, which included the same mCTX samples used to assess PHF1 as well as 4 additional ALS mCTX samples, using IHC. As described above, thionin was used to visualize the cells and the same AD samples were included as positive controls. pTau-S396 immunosignal was detected in both AD, ALS and control cases. As observed for PHF1 staining, extensive neuropil threads and NFTs were observed in AD brains (Figure 2P). Also, similar to PHF1, pTau-S396 immunosignal was heterogenous in control (Figure 2Q and R) and ALS mCTX (Figure 2S and T), therefore, the signal intensity was also categorized as outlined above for PHF1 in Table 4. Intense (+++) pTau-S396 immunostaining was detected in 22.2% of ALS and in 37.5% of control cases. Moderate (++) pTau-S396 staining was observed in 38.9% of ALS and 50% of controls, and weak (+) pTau-S396 staining was detected in 33.3% of ALS cases only. Extensive (+++) neuropil threads were detected in 27.8% of ALS and 12.5% of controls, moderate (++) neuropil threads in 27.8% of ALS and 75% of controls, and weak (+) neuropil threads in 38.9% of ALS cases. A high number (+++) of NFTs were detected in 5.6% of ALS cases, moderate number (++) of NFTs in 16.7% of ALS and 12.5% of controls, and 33.3% of ALS and 25% of control mCTX exhibited sparse NFTs (+). Lastly, pTau-S396 neuropil threads and staining were absent (-) in 5.6% of ALS and 12.5% of control mCTX, and no NFTs (-) were detected in 44.4% of ALS and in 62.5% of control cases (Table 4). In addition, the statistical analysis of the semi-quantitative immunosignal scores demonstrated that while there were no significant differences in pTau-S396 overall staining, and NFTs between ALS and controls (Fisher’s test, p = 0.2967, and p = 0.8975, respectively), there was a significant increase in pTau-S396 staining in ALS compared to control mCTX (Fisher’s test, p = 0.04627).

**Table 4.** pTau-S396 immunostaining in control and ALS post-mortem mCTX.

|  | Diffuse staining | Neuropil threads | NFTs | WM staining |
| --- | --- | --- | --- | --- |
| Control 1 | ++ | ++ | + | +++ |
| Control 2 | +++ | ++ | - | ++ |
| Control 3 | +++ | ++ | ++ | ++ |
| Control 4 | +++ | +++ | - | +++ |
| Control 5 | ++ | ++ | - | ++ |
| Control 6 | ++ | ++ | - | + |
| Control 7 | - | - | - | - |
| Control 8 | ++ | ++ | + | + |
| ALS 1 | ++ | + | - | + |
| ALS 2 | ++ | ++ | + | ++ |
| ALS 3 | +++ | +++ | + | +++ |
| ALS 4 | +++ | +++ | - | +++ |
| ALS 5 | ++ | ++ | + | ++ |
| ALS 6 | - | - | - | - |
| ALS 7 | + | + | ++ | +++ |
| ALS 8 | +++ | +++ | +++ | +++ |
| ALS 9 | +++ | +++ | + | +++ |
| ALS 10 | ++ | ++ | - | + |
| ALS 11 | + | ++ | ++ | + |
| ALS 12 | ++ | +++ | - | ++ |
| ALS 13 | ++ | ++ | + | ++ |
| ALS 14 | + | + | - | - |
| ALS 15 | + | + | ++ | - |
| ALS 16 | ++ | + | - | + |
| ALS 17 | + | + | - | - |
| ALS 18 | + | + | + | + |

Immunohistochemical pTau-S396 staining summary for diffuse staining, neuropil threads, NFTs and WM staining is given and indicated as follows: – no staining; + weak staining; ++ moderate staining; +++ intense staining.

Lastly, we assessed pTau levels in white matter (WM) in the same ALS and control mCTX samples to determine the levels of PHF1 and pTau-S396 staining and accumulation by IHC. As previously described, thionin was used to visualize the cells (Figure 3A-D). Similar to the staining in the gray matter, a heterogeneous PHF1 immunostaining was detected in both control (Figure 3E and F) and ALS mCTX (Figure 3G and H), therefore we categorized the intensity of the immunosignal as outlined in Table 3. Specifically, intense (+++) neuropil threads were detected in 7.1% of ALS and 12.5% of control mCTX, while moderate (++) neuropil threads were observed in 14.3% of ALS and 12.5% of control cases, and weak (+) neuropil threads were detected in 14.3% of ALS mCTX and 12.5% of control cases. PHF1 neuropil threads and staining were absent in 64.3% of ALS as well as in 62.5% of control cases (Table 3). The comparative semi-quantitative analysis using immunosignal scores revealed no significant change in WM PHF1 staining between control and ALS mCTX (Fisher’s test, p = 1). Similarly, pTau-S396 immunostaining in mCTX WM from control (Figure 3I and J) and ALS (Figure 3K and L) was categorized as outlined in Table 4. Intense (+++) neuropil threads were detected in 27.8% of ALS and 25% of control cases, moderate (++) neuropil threads in 22.2% of ALS and 37.5% of control mCTX, and weak (+) neuropil threads in 27.8% of ALS and 25% of control samples. Absence of neuropil threads was detected in 22.2% of ALS and 12.5% of control cases (Table 4). The statistical analysis of the semi-quantitative immunosignal scores demonstrated that there was no significant difference in WM pTau-S396 levels between ALS and control mCTX (Fisher’s test, p = 0.9408).

**Figure 3.**
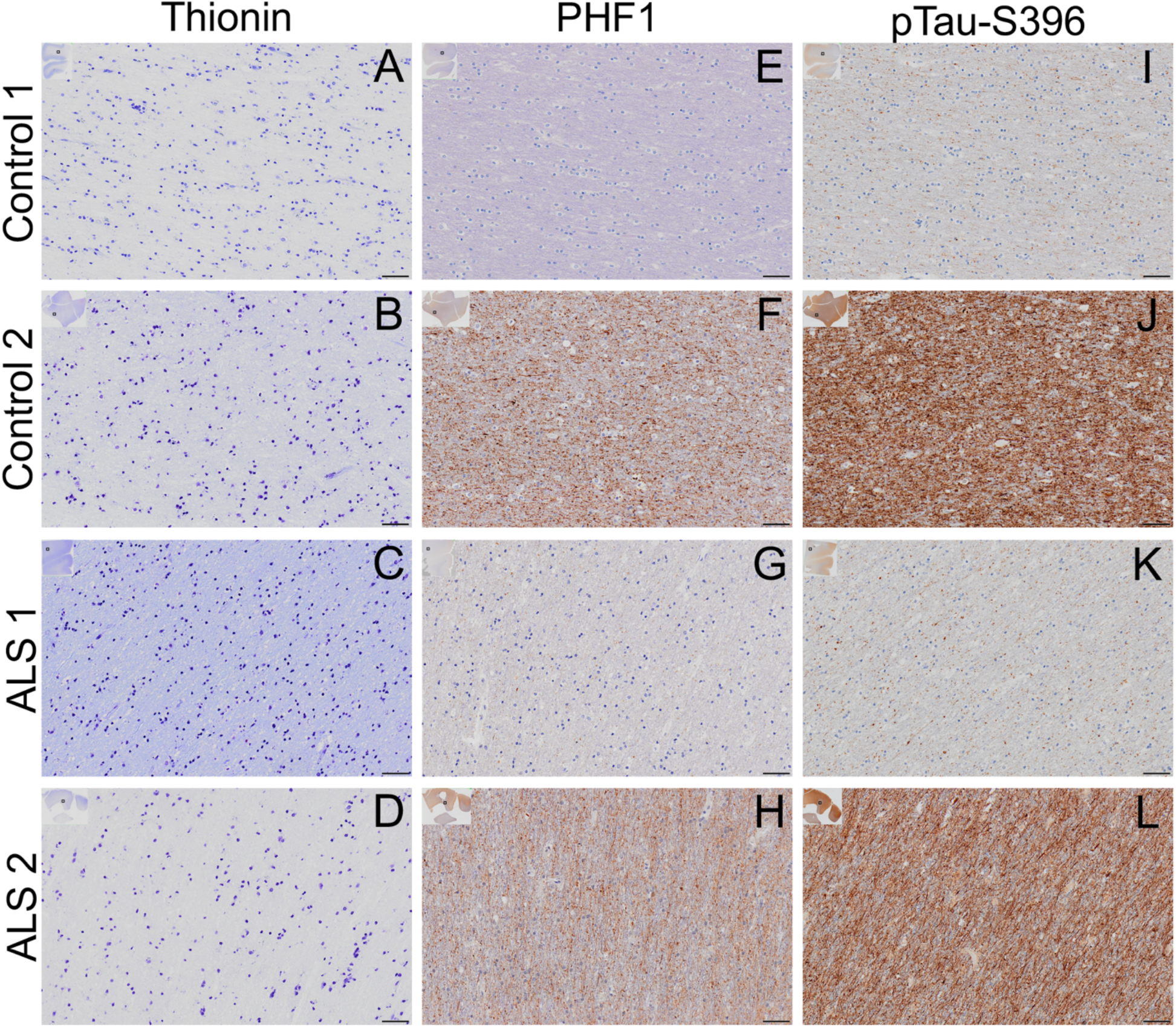
pTau accumulated in WM in ALS mCTX. (**A-D**) Thionin immunostaining in control and ALS post-mortem mCTX WM. Representative images of weak (+) PHF1 immunostaining in control mCTX (**E**) and ALS (**G**) WM. Representative images of intense (+++) PHF1 immunostaining in control (**F**) and ALS (**H**) WM. Representative images of weak (+) pTau-S396 immunostaining in control (**I**) and ALS (**K**) WM. Representative images of intense (+++) pTau-S396 immunostaining in control mCTX (**J**) and ALS (**L**) WM. Control (n = 8), ALS for PHF1 (n = 14), and ALS for pTau-S396 (n =18). Scale bar: 50 μM.

### Hyperphosphorylated tau mis-localizes to the nucleus in ALS post-mortem mCTX

Immunohistochemistry results also demonstrated an increase in both PHF1 and pTau-S396 immunostaining in the nucleus in ALS and control mCTX samples (Figure 4A-F). Therefore, we next assessed tau, pTau-S396, pTau-S404, and pTau-T181 levels in cytosolic and nuclear fractions derived from control (n = 15) and ALS (n = 25) mCTX. As shown in Figure 4G, histone H3 was only detected in the nuclear fractions from both control and ALS mCTX demonstrating a successful fractionation. While pTau-S396 and pTau-S404 levels were increased in the nuclear fraction in both ALS and control mCTX, tau and pTau-T181 levels were not changed. Two-way ANOVA demonstrated a significant effect of cellular fractions [F(1,76) = 8.080, p = 0.0057] with no significant effect of disease [F(1,76) = 0.4044, p = 0.5267] or cellular fractions X disease interaction [F(1,76) = 0.005833, p = 0.9393] on tau levels. Additionally, Tukey’s test revealed no change in cytosolic tau levels in ALS compared to the nuclear fraction (p = 0.9580) (Figure 4H). Similarly, two-way ANOVA demonstrated that, while there was a significant effect of cellular fractions [F(1,70) = 21.53, p < 0.0001], there was no significant effect of disease [F(1,70) = 0.7929, p = 0.3763] or cellular fractions X disease interaction [F(1,70) = 0.7633, p = 0.3853] on pTau-S396 levels. However, Tukey’s post-hoc analysis revealed a significant shift of pTau-S396 from the cytosolic to the nuclear fraction in ALS mCTX (p = 0.0002) (Figure 4I). In addition, two-way ANOVA demonstrated a significant effect of both cellular fractions [F(1,71) = 13.94, p = 0.0004] and disease [F(1,71) = 4.579, p = 0.0358], while there was no significant effect of cellular fractions X disease interaction [F(1,71) = 0.6897, p = 0.4090] on pTau-S404 levels. Similar to pTau-S396, Tukey’s post-hoc analysis revealed a significant shift of pTau-S404 from the cytosol to the nuclear fraction in ALS mCTX (p = 0.0017) (Figure 4J). Lastly, two-way ANOVA demonstrated that there was no significant effect of cellular fractions [F(1,73) = 0.05855, p = 0.8095], disease [F(1,73) = 1.580, p = 0.2127], or cellular fractions X disease interaction [F(1,73) = 0.02573, p = 0.8730] on pTau-T181 levels and Tukey’s post-hoc analysis revealed no change in pTau-T181 levels in ALS nuclear fraction compared to the cytosolic fraction (p = 0.9877) (Figure 4K).

**Figure 4.**
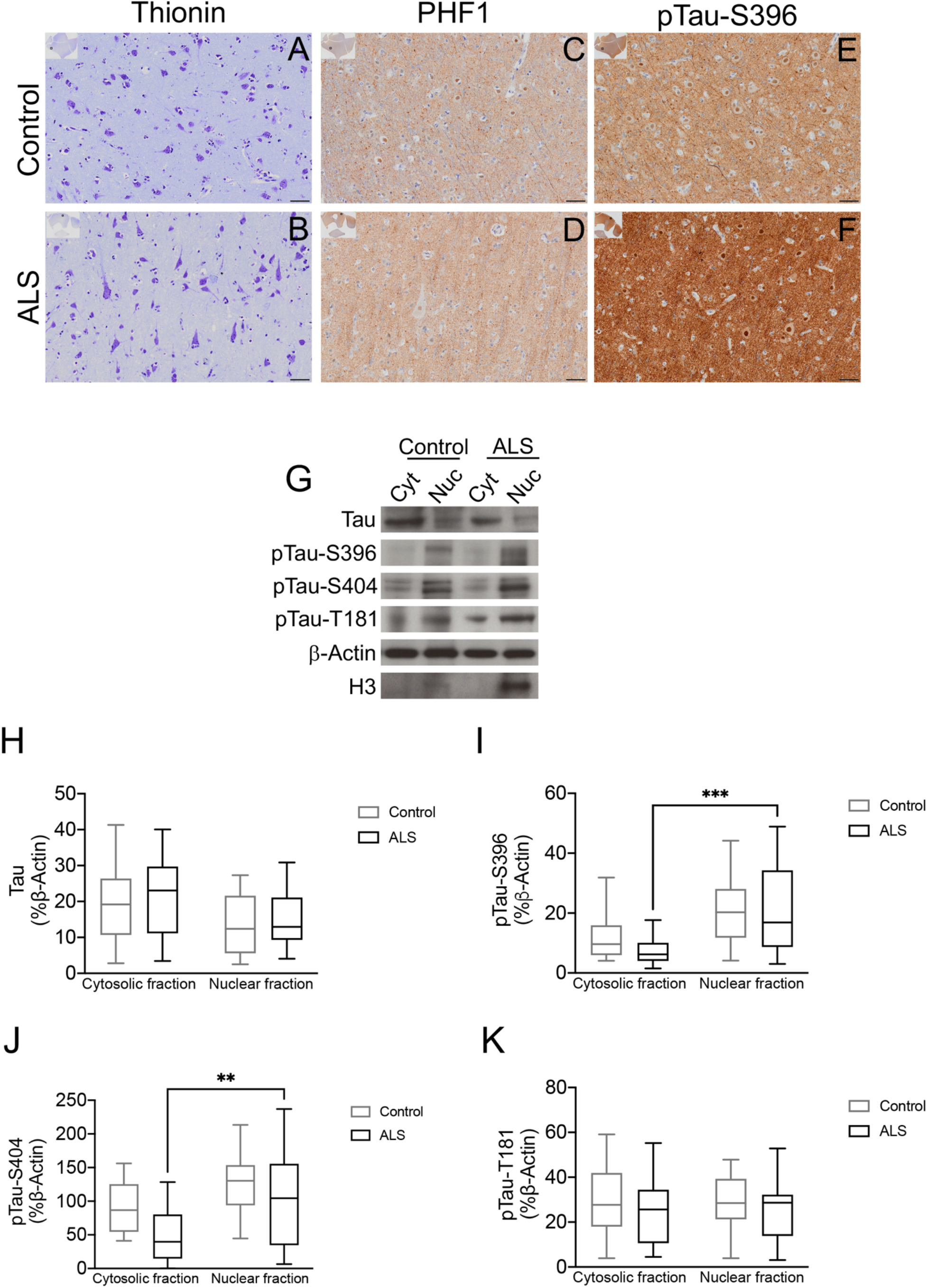
Hyperphosphorylated tau is mis-localized to the nucleus in ALS mCTX. Thionin immunostaining in control (**A**) and ALS (**B**) mCTX. PHF1 immunostaining in control (**C**) and ALS (**D**) mCTX demonstrating nuclear pTau. pTau-S396 immunostaining in control (**E**) and ALS (**F**) mCTX demonstrating nuclear pTau. Representative western blot images of tau, pTau-S396, pTau-S404, pTau-T181, and histone H3 in cytosolic and nuclear fractions from control and ALS mCTX **(G)**. There was no significant difference in tau levels between nuclear and cytosolic fractions in ALS (Tukey’s test, p = 0.9580) (**H**). pTau-S396 levels were significantly increased in nuclear compared to cytosolic fractions in ALS (Tukey’s test, p = 0.0002) (**I**). pTau-S404 levels were significantly increased in nuclear compared to cytosolic fractions in ALS (Tukey’s test, p = 0.0017) (**J**). There was no change in pTau-T181 levels in ALS nuclear or cytosolic fractions (Tukey’s test, p = 0.9877) (**K**). For IHC Control (n = 8), ALS for PHF1 (n = 14), and ALS for pTau-S396 (n =18). The graphs demonstrate box plots with the central line representing the median, the edges representing the interquartile range, and the whiskers representing the minimum and maximum values from western blot experiments performed in control (n = 15) and ALS mCTX (n = 25) following cytosolic/nuclear fractionation. Scale bar: 50 μM. ** p < 0.01; *** p < 0.001.

### Nuclear pTau is increased in ALS independent of sex, region of onset, and genotype

Next, we correlated increases in pTau at the nuclear level in ALS with the known patient clinical information when available. The analysis revealed a significant shift in pTau-S396 and pTau-S404 from the cytosolic fraction to the nuclear fraction in male (n = 11) (Mann Whitney U = 18, p = 0.0079, and Mann Whitney U = 24, p = 0.0158, respectively) and female ALS mCTX (n = 14) (Mann Whitney U = 29.50, p = 0.0037, and Mann Whitney U = 38, p = 0.0091, respectively) (Suppl. Figure 1A-D). While pTau-S396 levels were increased in the nuclear fraction compared to the cytosolic fraction in bulbar ALS (n = 10), there was no change in pTau-S404 (Mann Whitney U = 17, p = 0.0379, and Mann Whitney U = 27, p = 0.1564) (Suppl. Figure 1E and F). However, both pTau-S396 and pTau-S404 levels were significantly increased in the nuclear fraction compared to the cytosolic fraction in limb onset ALS (n = 12) (Mann Whitney U = 30, p = 0.0257, and Mann Whitney U = 22, p = 0.0104, respectively) (Suppl. Figure 1G and H). While there were no significant difference in pTau-S396 and pTau-S404 in the nuclear fraction compared to cytosolic fractions in ALS/FTD (n = 3) (Mann Whitney U = 3, p = 0.7000, and Mann Whitney U = 2, p = 0.4000, respectively) (Suppl. Figure 1I and J), there was a significant increase in nuclear pTau-S396 and pTau-S404 in ALS cases associated with TDP-43 proteinopathy (n = 14) (Mann Whitney U = 29, p = 0.0019, and Mann Whitney U = 36, p = 0.0066, respectively) (Suppl. Figure 1K and L). Lastly, while there was a trend towards a significant shift in pTau-S396 levels from the cytosolic fraction to the nuclear fraction in C9ORF72-ALS (n = 5), there were no alterations in pTau-S404 levels (Mann Whitney U = 4, p = 0.0952, and Mann Whitney U = 8, p = 0.4206, respectively) (Suppl. Figure 1M and N). The same shift was also observed in the single SOD1-ALS case used in this study (Suppl. Figure 1O and P).

### Hyperphosphorylated tau mis-localizes to synapses in ALS

Given that hyperphosphorylated tau accumulates at the synaptic level in Alzheimer’s disease (AD) [34], we assessed the levels of tau, and its phosphorylated isoforms in three different mCTX fractions – total, cytosolic, and SNs fractions – from ALS (n = 36) and non-neurological control (n = 14) by western blots. As demonstrated in Figure 5A, while PSD-95 was detected in the total and SNs fractions, it was absent in the cytosolic fraction derived from both ALS and control mCTX, demonstrating successful fractionation and purity of SNs. Western blot analysis revealed that tau, pTau-S396, and pTau-S404 shift to SNs in ALS mCTX. Specifically, when assessing tau levels, two-way ANOVA demonstrated a significant effect of cellular fractions [F(2, 137) = 15.03, p < 0.0001], with no effect of disease [F(1,137) = 0.1050, p = 0.7464], or cellular fractions X disease interaction [F(2,137) = 0.3903, p = 0.6776]. Furthermore, Tukey’s post-hoc analysis revealed a significant increase in tau levels in SNs compared to total extract in ALS mCTX (p = 0.0489). There was also a significant increase in tau levels in both control- and ALS-derived SNs compared to their cytosolic fractions (p = 0.0059, and p = 0.0006, respectively) (Figure 5B). Assessment of pTau-S396 levels demonstrated that while there was a significant effect of cellular fractions [F(2,133) = 4.106, p = 0.0186], there was no significant effect of disease [F(2,133) = 1.320, p = 0.2527], or cellular fractions X disease interaction [F(2,133) = 0.9085, p = 0.4056] as measured by two-way ANOVA. Moreover, Tukey’s post-hoc analysis revealed a significant shift of pTau-S396 from ALS cytosolic fraction to the SNs (p = 0.0024) (Figure 5C). Similarly, two-way ANOVA demonstrated a significant effect of cellular fractions [F(2,131) = 6.701, p = 0.0017] on pTau-S404 levels. However, there were no significant effects of disease [F(1,131) = 0.04490, p = 0.8325] or cellular fractions X disease interaction [F(2,131) = 0.7333, p = 0.4823] on pTau-S404 levels. Tukey’s post-hoc analysis revealed a significant decrease in pTau-S404 levels in ALS-derived cytosolic fraction compared to the total extract (p = 0.0019) and a significant shift of pTau-S404 from the cytosol to the SNs in ALS mCTX (p = 0.0016) (Figure 5D). Lastly, assessment pTau-T181 levels using two-way ANOVA revealed a significant effect of cellular fractions [F(1,130) = 3.526, p = 0.0323], disease [F(1,130) = 4.296, p = 0.0402], and cellular fractions X disease interaction [F(2,130) = 3.626, p = 0.0294] and Tukey’s post-hoc analysis demonstrated a significant decrease in pTau-T181 levels in ALS-derived SNs compared to controls (p = 0.0099) (Figure 5E).

**Figure 5.**
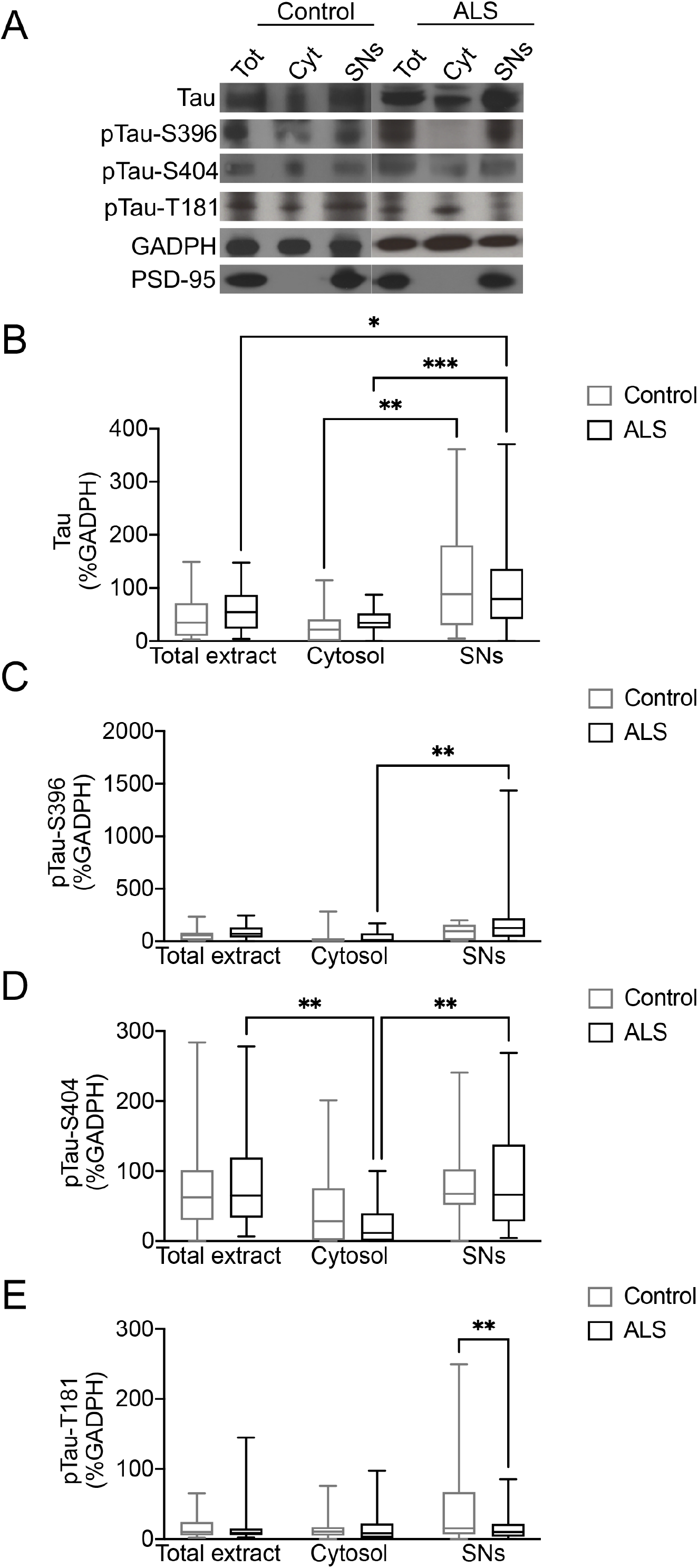
pTau-S396 and pTau-S404 are increased in ALS SNs. (**A**) Representative western blot images of tau, pTau-S396, pTau-S404, pTau-T181, and PSD-95 in total, cytosolic, and SNs fractions derived from control and ALS mCTX. **(B)** There was a significant increase in tau levels in control- and ALS-derived SNs compared to the cytosolic fractions (Tukey’s test, p = 0.0059, and p = 0.0006, respectively). **(C)** pTau-S396 levels were significantly increased in ALS SNs compared to cytosolic fraction (Tukey’s test, p = 0.0024). **(D)** There was a significant decrease in pTau-S404 levels in ALS cytosolic fraction compared to total extract and SNs (Tukey’s test, p = 0.0019, and p = 0.0016, respectively). **(E)** pTau-T181 levels were significantly decreased in ALS SNs compared to control SNs (Tukey’s test, p = 0.0099). The graphs demonstrate box plots with the central line representing the median, the edges representing the interquartile ranges, and the whiskers representing the minimum and maximum values from western blot experiments performed in control (n = 14) and ALS mCTX (n = 36) following SNs fractionation. * p < 0.05; ** p < 0.01; *** p < 0.001.

### pTau-S396 and pTau-S404 are increased in ALS SNs independent of sex, region of onset, and genotype

Next, we correlated increases in pTau in ALS SNs with the known patient clinical information when available. The analysis revealed a significant shift in pTau-S396 and pTau-S404 from the cytosolic fraction to the SNs in male (n = 20) (Mann Whitney U = 49, p = 0.0004, and Mann Whitney U = 46, p = 0.0014, respectively) and female ALS patients (n = 16) (Mann Whitney U = 42, p = 0.0026, and Mann Whitney U = 19, p < 0.0001, respectively) (Suppl. Figure 2A-D). Furthermore, pTau-S396 and pTau-S404 levels were increased in the SNs compared to the corresponding cytosolic fraction in both bulbar onset (n = 13) (Mann Whitney U = 21, p = 0.0023, and Mann Whitney U = 5.500, p < 0.0001, respectively) and limb onset ALS (n = 19) (Mann Whitney U = 58, p = 0.0077, and Mann Whitney U = 50.50, p = 0.0089, respectively) (Suppl. Figure 2E-H). While there were no significant difference in pTau-S396 and pTau-S404 in SNs compared to the cytosolic fraction in ALS/FTD (n = 3) (Mann Whitney U = 0, p = 0.1000, and Mann Whitney U = 0, p = 0.1000, respectively) (Suppl. Figure 2I and J), there was a significant increase in both pTau-S396 and pTau-S404 in SNs in ALS cases with TDP-43 proteinopathy (n = 18) (Mann Whitney U = 46, p = 0.0008, and Mann Whitney U = 47.50, p = 0.0018, respectively) (Suppl. Figure 2K and L). Lastly, there was a significant shift in both pTau-S396 and pTau-S404 from the cytosol to the SNs in C9ORF72-ALS (n = 5) (Mann Whitney U = 1, p = 0.0159, and Mann Whitney U = 2, p = 0.0317, respectively) and the same shift was observed in the single SOD1-ALS case used in this study (Suppl. Figure 2M-P).

### pTau-T181:tau ratio is decreased in ALS CSF

Recent studies have suggested that alterations in tau and pTau-T181 may be viable biomarkers for AD [38-41], therefore, we assessed the levels of total tau and pTau-T181 in CSF from healthy controls (n = 10) and ALS patients (n = 40) using Quanterix Simoa assays. The analysis indicated that there was no significant difference in total tau (Mann Whitney U test = 100, p = 0.1234) or pTau-T181 levels (Mann Whitney U test = 147, p = 0.8227) in ALS CSF compared to control CSF (Figure 6A and B). Importantly, there was a significant decrease in pTau-T181:tau ratio in ALS CSF compared to control samples (Mann Whitney U test = 56, p = 0.0025) (Figure 6C). Next, we correlated alterations in tau and pTau in ALS CSF with the known patient clinical information when available. The analysis revealed that while CSF tau levels were significantly increased in ALS patients diagnosed with bulbar onset disease (n = 6) (Mann Whitney U = 9, p = 0.0225), there was a significant decrease in pTau-T181:tau ratio (Mann Whitney U = 7, p = 0.0110) and no change in CSF pTau-T181 levels (Mann Whitney U = 27, p = 0.7925) in this group of patients (Figure 6D-F). Similarly, pTau-T181:tau ratio was significantly decreased in CSF from limb onset ALS patients (n = 25) (Mann Whitney U = 49, = 0.0045) while tau and pTau-T181 levels were not altered in this group (Mann Whitney U = 91, p = 0.2253, and Mann Whitney U = 123, p = 0.8214, respectively) (Figure 6G-I).

**Figure 6.**
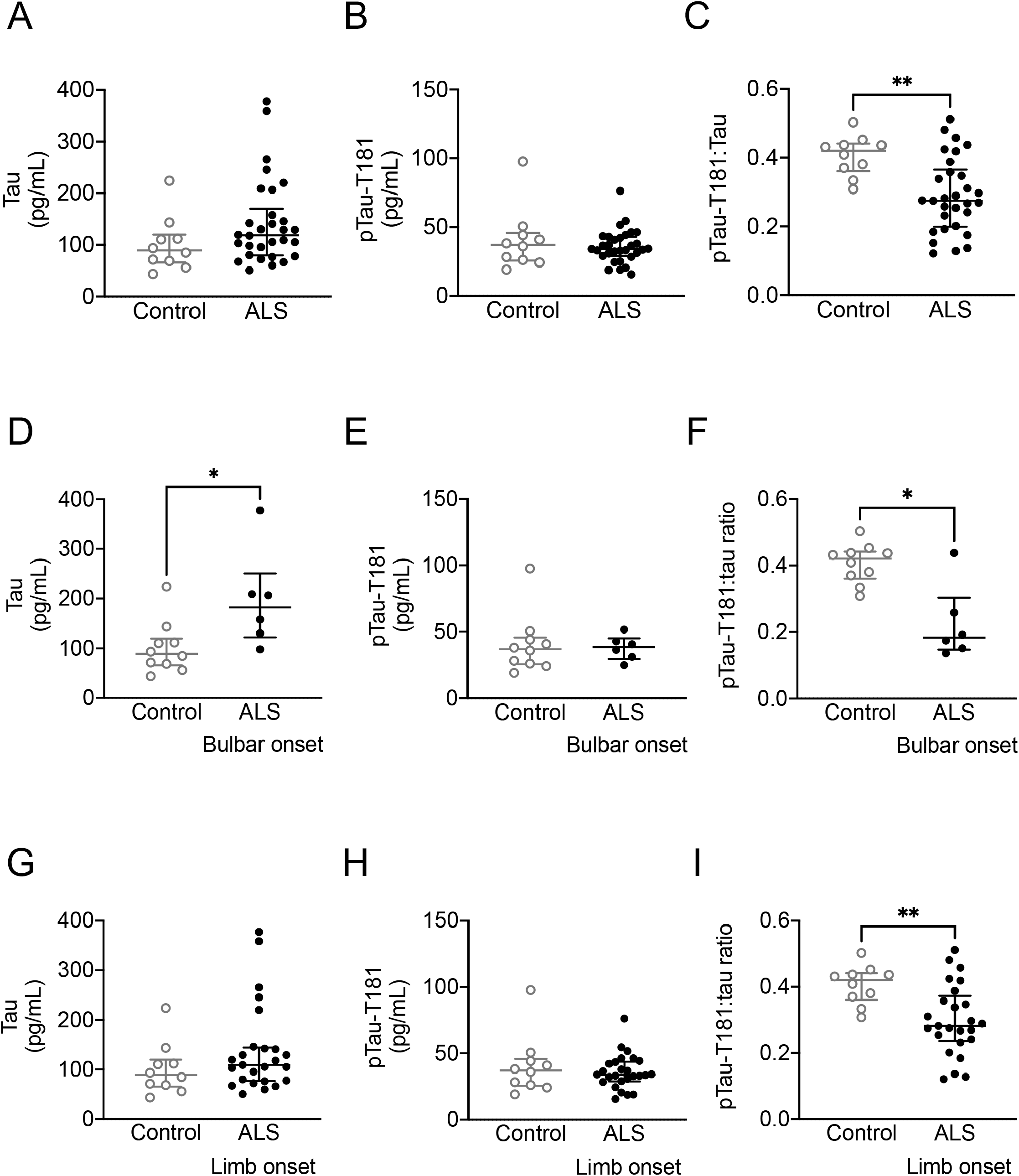
CSF pTau-T181:tau ratio is decreased in ALS. **(A)** There was no difference in CSF tau levels between ALS patients and healthy controls (Mann Whitney U test = 100, p = 0.1234). **(B)** CSF pTau-T181 levels were not different between ALS and controls (Mann Whitney U test = 147, p = 0.8227). **(C)** There was a significant decrease in pTau-T181:tau ratio in ALS CSF compared to healthy controls (Mann Whitney U test = 56, p = 0.0025). **(D)** There was a significant increase in CSF tau levels in bulbar onset ALS (n = 6) compared to controls (Mann Whitney U = 9, p = 0.0225). (**E**) CSF pTau-T181 levels were not altered in bulbar onset ALS (n = 6) (Mann Whitney U = 27, p = 0.7925). (**F**) There was a significant decrease in CSF pTau-T181:tau ratio in bulbar onset ALS (n = 6) (Mann Whitney U = 7, p = 0.0110). (**G**) There was no difference in CSF tau levels in limb onset ALS (n = 25) (Mann Whitney U = 91, p = 0.2253). (**H**) CSF pTau-T181 levels were not altered in limb onset ALS (n = 25) (Mann Whitney U = 123, p = 0.8214). (**I**) There was a significant decrease in CSF pTau-T181:tau ratio in limb onset ALS (n = 25) (Mann Whitney U = 49, = 0.0045). The graphs demonstrate the individual values of tau, pTau-T181, and pTau-T181:tau ratio from Quanterix Simoa Assay performed in CSF from ALS (n = 40) and non-neurological control (n = 10), with the central line representing the median, and the edges representing the interquartile range. * p < 0.05; ** p < 0.01.

Further analysis revealed that there was no correlation between tau, pTau-T181, or pTau-T181:tau ratio and the age of patients at time of sample collection (Spearman correlation, p = 0.376, p = 0.442, and p = 0.736 respectively) (Figure 7A-C). Similarly, there was no correlation between tau or pTau-T181 levels and disease duration (Spearman correlation, p = 0.835 and p = 0.261, respectively) (Figure 7D and E), however there was a trend towards a significant correlation between pTau-T181:tau ratio and disease duration (Spearman correlation, p = 0.063) (Figure 7F). There was also a trend towards a significant correlation between the ALSFRS-R at time of first visit with tau levels (Spearman correlation, p = 0.057), however, ALSFRS-R did not correlate with pTau-T181 or pTau-T181:tau ratio levels (Spearman correlation, p = 0.546 and p = 0.222, respectively) (Figure 7G-I). Furthermore, while there was no correlation between pTau-T181 levels and pre-baseline ALSFRS-R slope (Spearman correlation, p = 0.494, respectively), there was a trend towards a significant correlation between tau levels and pre-baseline ALSFRS-R slope (Spearman correlation, p = 0.057) as well as a significant correlation between pre-baseline ALSFRS-R slope and pTau-T181:tau ratio (Spearman correlation, p = 0.005) (Figure 7J-L).

**Figure 7.**
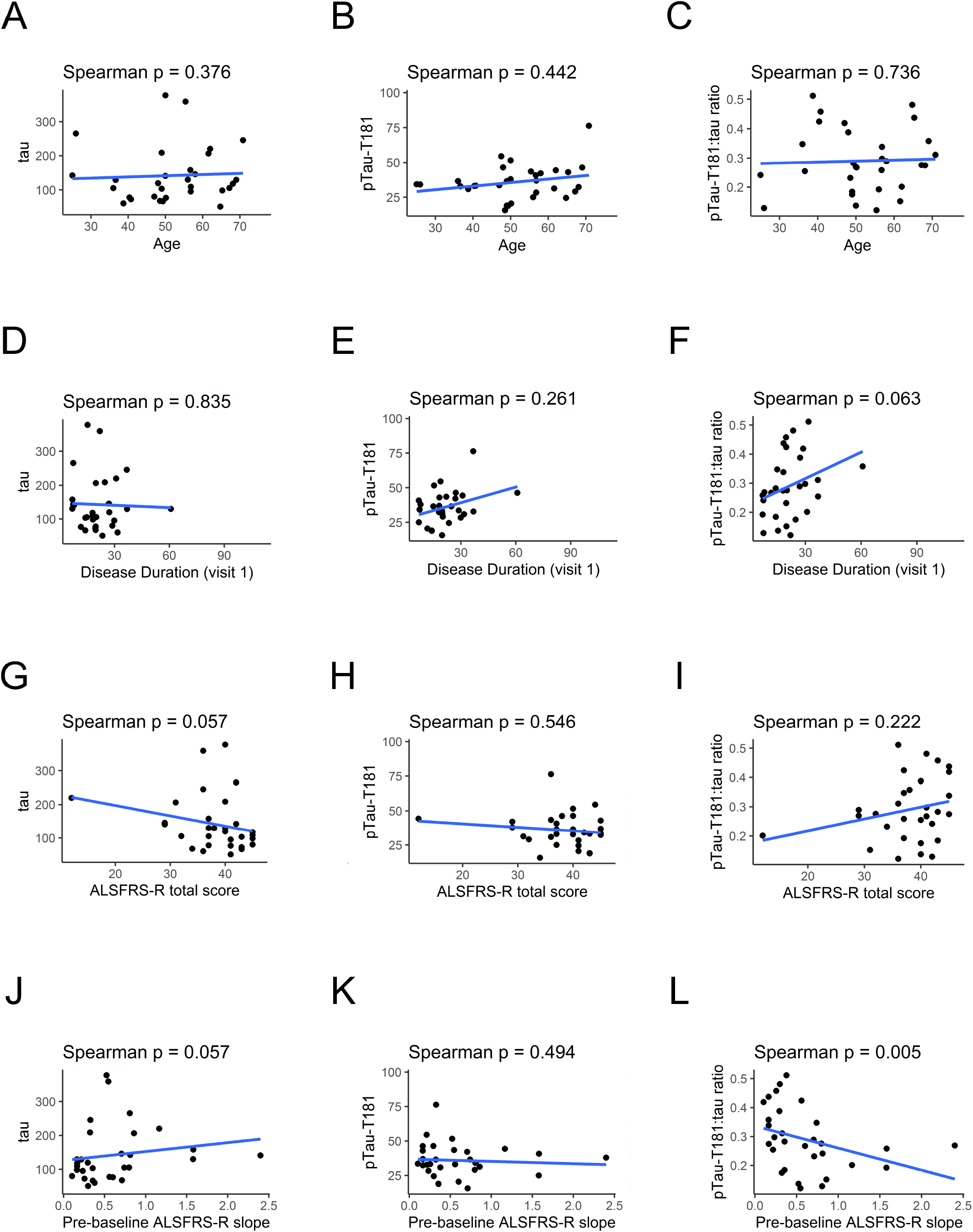
CSF Tau and pTau-T181:tau ratio correlate with ALSFRS-R. (**A**) There was no correlation between CSF tau levels and age of ALS patients at the first visit (Spearman correlation, p = 0.376). (**B**) CSF pTau-T181 levels did not correlate with age of ALS patients at the first visit (Spearman correlation, p = 0.442). (**C**) There was no correlation between pTau-T181:tau ratio and age of ALS patients at the first visit (Spearman correlation, p = 0.736). (**D**) CSF tau levels did not correlate with ALS disease duration (Spearman correlation, p = 0.835). (**E**) There was no correlation between CSF pTau-T181 levels and disease duration (Spearman test, p = 0.261). (**F**) There was a trend towards a significant correlation between CSF pTau-T181:tau ratio and ALS disease duration (Spearman correlation, p = 0.063). (**G**) There was a trend towards a significant inverse correlation between CSF tau levels and ALSFRS-R (Spearman correlation, p = 0.057). (**H**) There was no correlation between CSF pTau-T181 levels and ALSFRS-R (Spearman correlation, p = 0.546). (**I**) CSF pTau-T181:tau ratio did not correlate with ALSFRS-R (Spearman correlation, p = 0.222). (**J**) There was a trend towards a negative correlation between CSF tau levels and ALSFRS-R slope (Spearman correlation, p = 0.057). (**K**) CSF pTau-T181 levels did not correlate with ALSFRS-R slope (Spearman correlation, p = 0.494). (**L**) There was a positive correlation between CSF pTau-T181:tau ratio and ALSFRS-R slope (Spearman correlation, p = 0.005). The graphs represent correlations between CSF tau, pTau-T181, or pTau-T181:tau ratio and age, disease duration, ALSFRS-R or ALSFRS-R slope.

Lastly, longitudinal analysis of ALS patient CSF revealed that there was high variability in both tau and pTau-T181 trajectory. Specifically, the analysis of trajectories revealed that while there was a significant decline over time based on the ALSFRS-R assessed for each patient at each visit (p < 0.001), there were not significant alterations in tau (p = 0.907), pTau-T181 (p = 0.222), or pTau-T181:tau ratio in ALS (p = 0.578) over time (Figure 8A-D) (Table 5). Alterations in tau over an average of 11 months of follow-up demonstrated no significant correlation with change in ALSFRS-R (p = 0.662), pTau-T181 or pTau-T181:tau ratio (average follow-up 9 months; p=0.185 and p = 0.327, respectively; Supplementary Figure 3A-C; Supplementary Tables 1-3). When tau was added to the ALSFRS-R trajectory model, there was a significant effect of tau on the change in ALSFRS-R over time (p=0.024) (Table 6). This model estimated a baseline ALSFRS-R total score of 37.9 points and a slope of −0.56 point/month and a 10-point increase in tau at baseline resulted in a further −0.026 point/month change in ALSFRS-R. While pTau-T181 showed no significant effects (p = 0.240), pTau-T181:tau ratio revealed a trend towards a significant effect (p = 0.051) (Tables 7 and 8).

**Table 5.** Trajectories of ALSFRS-R, tau, pTau-T181, and pTau-T181: tau ratio in ALS CSF.

|  | Intercept | Slope | Slope SE | Slope p |
| --- | --- | --- | --- | --- |
| ALSFRS-R | 37.8 | -0.47 | 0.06 | < 0.001 |
| Tau | 147.9 | 0.16 | 1.38 | 0.907 |
| pTau-T181 | 39.07 | -0.09 | 0.07 | 0.222 |
| pTau-T181:tau ratio | 0.3 | 0 | 0 | 0.578 |

**Table 6.**
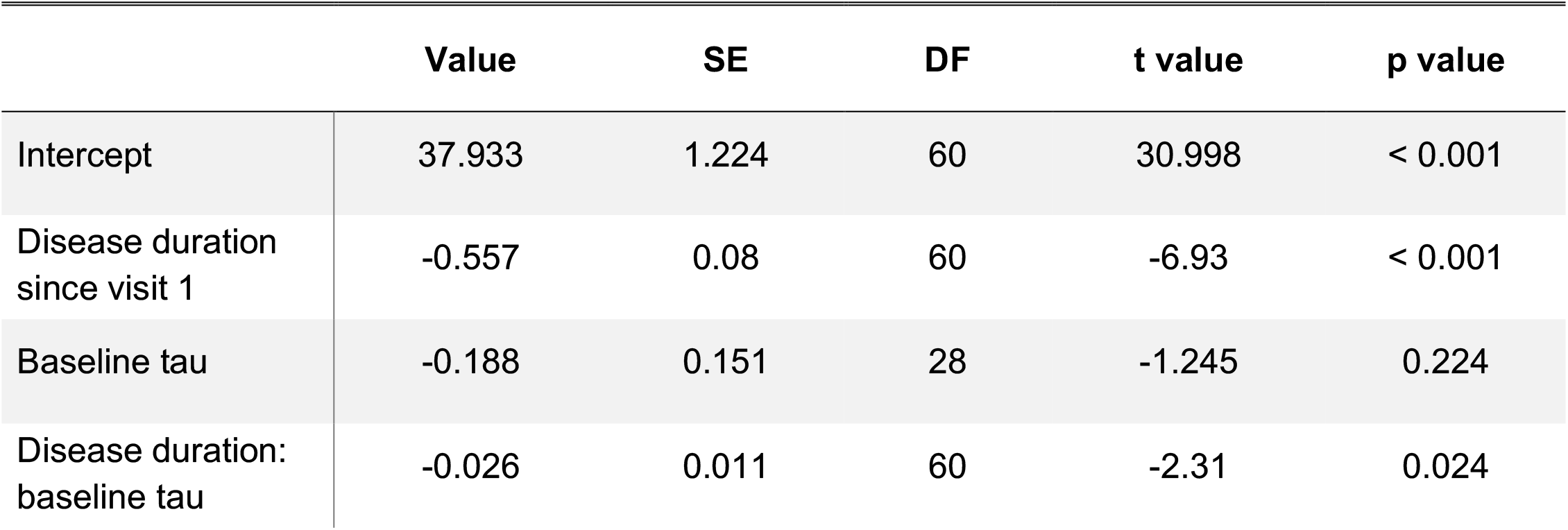
Effect of baseline tau on ALSFRS-R trajectory.

**Table 7.** Effect of baseline pTau-T181 on ALSFRS-R trajectory.

|  | Value | SE | DF | t value | p value |
| --- | --- | --- | --- | --- | --- |
| Intercept | 38.111 | 1.229 | 65 | 30.999 | < 0.001 |
| Disease duration since visit 1 | -0.493 | 0.075 | 65 | -6.608 | < 0.001 |
| Baseline pTau-T181 | -0.612 | 1.034 | 29 | -0.592 | 0.558 |
| Disease duration: baseline pTau-T181 | -0.066 | 0.055 | 65 | -1.186 | 0.240 |

**Table 8.** Effect of baseline pTau-T181:tau ratio on ALSFRS-R trajectory.

|  | Value | SE | DF | t value | p value |
| --- | --- | --- | --- | --- | --- |
| Intercept | 37.964 | 1.227 | 60 | 30.929 | < 0.001 |
| Disease duration since visit 1 | -0.571 | 0.09 | 60 | -6.36 | < 0.001 |
| Baseline pTau-T181:tau ratio | 13.89 | 11.492 | 28 | 1.209 | 0.237 |
| Disease duration: baseline pTau-T181:tau ratio | 1.423 | 0.715 | 60 | 1.989 | 0.051 |

**Figure 8.**
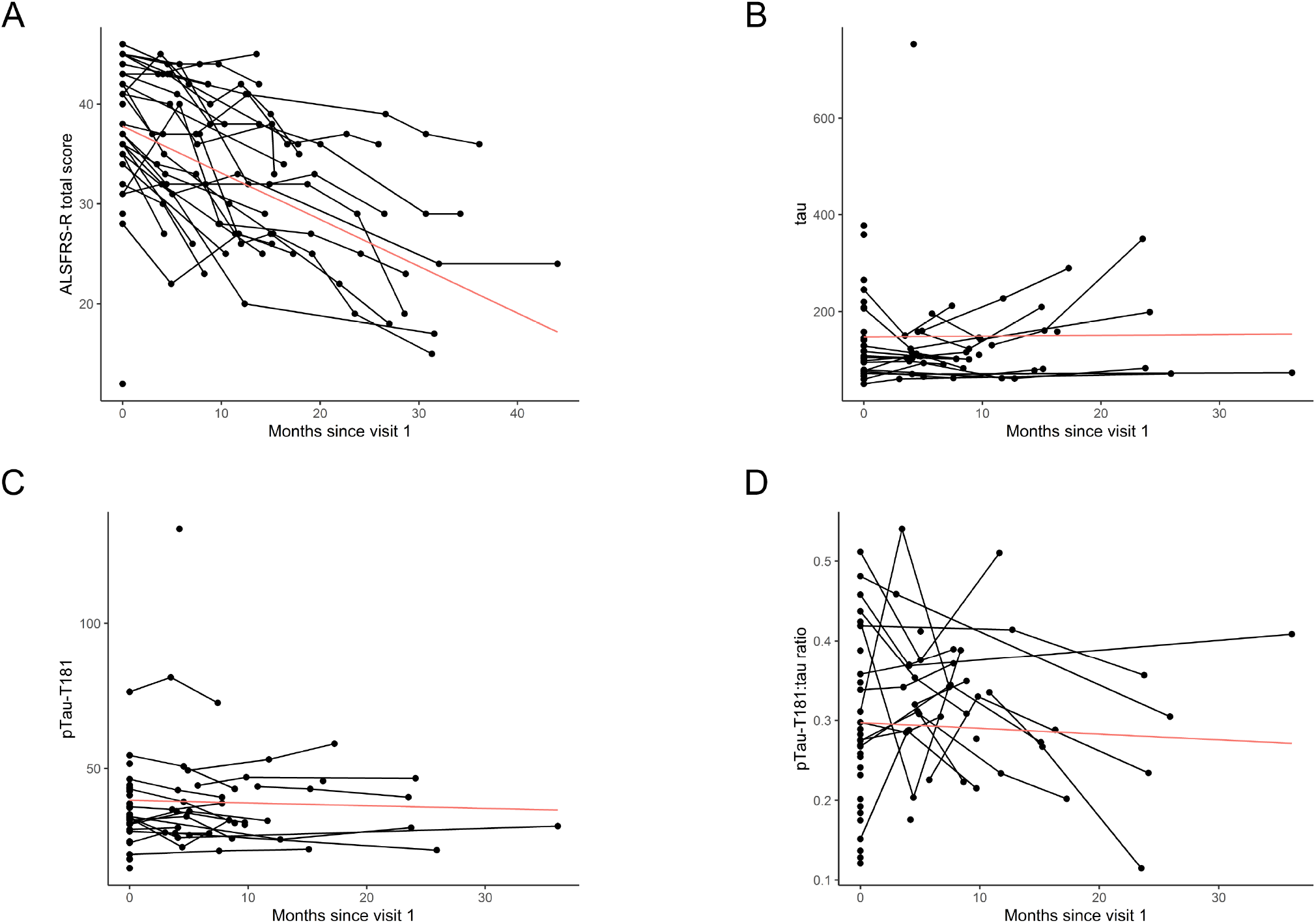
Tau levels and pTau-T181:tau ratio in CSF correlate with functional decline in ALS. (**A**) There was a significant decline in ALSFRS-R over time based on the ALSFRS-R scores assessed for each patient at different visits (p < 0.001). (**B**) There were no significant alterations in CSF tau levels (p = 0.907). (**C**) CSF pTau-T181 levels were not significantly altered (p = 0.222). (**D**) CSF pTau-T181:tau ratio was not significantly altered (p = 0.578). The graphs represent the trajectories of variables over time.

## Discussion

Our findings provide evidence that C-terminus phosphorylation of tau leads to its mis-localization to the nucleus and synapses in ALS mCTX and that decreases in pTau-T181:tau ratio in ALS CSF may be a viable biomarker for ALS. We also identified novel *MAPT* variants clustering near the C-terminus domain in ALS patients that were absent in non-ALS cases using the data from ALSKP browser and Project MinE. Furthermore, we demonstrated that although bulk tau phosphorylation was not altered in ALS mCTX, there was a significant increase in nuclear pTau-S396 and pTau-S404 levels in ALS. While pTau-S396 and pTau-S404 were shown to be mis-localized to synapses in ALS post-mortem mCTX, there was a decrease in pTau-T181 levels in ALS SNs compared to control SNs. Lastly, we report that total tau and pTau-T181 levels were not significantly altered in ALS CSF, however, there was a significant decrease in the pTau-T181:tau ratio in ALS CSF, as previously reported [22, 25, 42]. Interestingly, CSF tau levels were significantly increased in ALS patients diagnosed with bulbar onset, while pTau-T181:tau ratio was significantly decreased in both bulbar and limb onset ALS patients. Importantly, tau levels in ALS CSF were inversely correlated with the ALSFRS-R, while pTau-T181:tau ratio positively correlated with ALSFRS-R. While there were no alterations in tau, pTau-T181, and pTau-T181:tau ratio during disease progression, there was an increase in the rate of decline per month in ALS patients that correlated with increases in tau levels and was inversely correlated with the pTau-T181:tau ratio.

We identified 36 *MAPT* genetic variants in 42 ALS patients from ALSKP and Project MinE data browsers. Of the 36 variants, 15 were unique to ALS cases and the remaining 21 were observed at a very low frequency in gnomAD. Thirty-three of 36 were missense variants, which we subsequently annotated using CADD and MPC, and classified into 5 pathogenicity categories. Interestingly, the eight ALS variants associated with a high predicted pathogenicity cluster near the C-terminus of the protein transcript within or around the microtubule-binding domain consistent with the other pathogenic or likely pathogenic variants in patients with FTD and motor neuron dysfunction previously reported in ClinVar. Specifically, two variants, p.Leu583Val and p.Lys652Met, which were collectively observed in four unrelated ALS patients in ALSKP and Project MinE, were also observed in pedigrees displaying FTD, parkinsonism, and motor neuron degeneration. The recurrence of these variants may suggest either a common haplotype among the four ALS patients and these pedigrees or a mutation hotspot. Given the evidence of variant pathogenicity clustering within or neighboring the microtubule-binding domain in multiple pedigrees with neurodegeneration, it is likely these eight variants observed in ALS cases explain or significantly contribute to neurodegeneration in these patients. The molecular consequence of the remaining variants outside of the microtubule-binding domain is unclear and may not contribute to the development of ALS or other overlapping neurodegenerative disorders.

Additionally, our findings support and extend recent descriptions of tau pathology in both sporadic and familial forms of ALS [5-6] as well as in ALS/Parkinsonism dementia complex (ALS/PDC) [43-44]. Increases in total tau and cytoplasmic inclusions of hyperphosphorylated tau have been described in post-mortem mCTX and spinal cord from ALS patients [19-21]. However, our findings here demonstrate that there is not an overall increase in tau phosphorylation in ALS. Accordingly, tau immunostaining is heterogenous in post-mortem mCTX and tau neuropil threads and sparse NFTs are present in both ALS and controls. Similarly, we did not observe quantitative differences in tau or pTau levels in ALS mCTX compared to controls. Interestingly, tau neuropil threads were identified in ALS in the white matter, raising the question of whether this may underlie deficits in axonal transport, one of the suggested pathogenic mechanisms underlying motor neuron loss in ALS [45-50]. Future and ongoing studies are focused on delineating the pathogenic role of pTau accumulation in WM and how it may relate to axonal degeneration in ALS.

Our data also revealed an increase in two C-terminus pTaus (S396 and S404) at the nuclear level in ALS mCTX across subtypes of the disease, which could tie tau pathology to another key mechanism of pathogenesis in ALS, nucleocytoplasmic transport [27, 51-56]. Indeed, the impairment of nuclear transport leads to the cytoplasmic accumulation of nuclear proteins, including TDP-43, FUS, hnRNPA1, and MATR3 in ALS [57-60]. Specifically, hyperphosphorylation of tau favors tau interactions with nucleoporins and, in turn, triggers alterations in the nuclear pore complex [61]. However, future studies will be necessary to clarify the pathogenetic role of pTau accumulation at the nuclear level in ALS.

We also demonstrated mis-localization of pTau-S396 and pTau-S404 at the synaptic levels in ALS mCTX, reminiscent of AD [34]. Interestingly, pTau-S396 and pTau-S404 accumulate in the SNs across subtypes of the disease, thus suggesting that pTau mis-localization at the synaptic level may represent a common feature underlying ALS pathogenesis, although additional studies with a larger sample size are required to determine the pathology of genetic subtypes. Of note, while there is an overall increase in tau phosphorylation in AD and in other tauopathies, we reported a decrease at the N-terminus phosphorylation at T181 in ALS SNs, thus suggesting a specific hyperphosphorylation of the C-terminus domain in ALS. Furthermore, it is possible that the decrease in pTau-T181 levels at the synaptic level may suggest a protective role for tau phosphorylation at the N-terminus in ALS. This notion is supported by our CSF data indicating a decrease in the rate of decline per month in response to increases in pTau-T181 levels in relation to total tau levels. However, further investigation focused on the pathogenic role of different phosphorylation sites at both the C- and N-terminus are necessary to validate this hypothesis.

Recent studies suggest that alterations in tau levels in patient biofluids, such as CSF, may serve as a biomarker for the diagnosis of tauopathies [62-64]. Specifically, CSF tau and pTau-T181 have been suggested as biomarkers for AD given that levels of both are increased in patients [38-41]. Whether tau levels could serve as a viable biomarker in ALS remains contradictory in the field as the results vary between each study. To date, several studies have assessed CSF tau in ALS patients demonstrating both an increase and no change in tau in ALS [22-25, 42, 65]. While all studies demonstrate no significant difference in pTau-T181 levels in ALS CSF, several reports demonstrate significant decreases in pTau:tau ratio in ALS CSF [22, 25, 66]. Similarly, controversial data have been reported regarding the potential prognostic validity of CSF tau in ALS [67-70], while other studies suggest a positive correlation between tau and pTau levels at baseline and disease progression [22, 71] as well as a correlation between pTau:tau ratio and disease progression [25]. Our findings here demonstrate that while there are no alterations in tau and pTau-T181 in ALS CSF, there is a significant decrease in the pTau-T181:tau ratio, supporting previously published reports [22, 25, 42, 66]. Importantly, increases in CSF tau levels correlate with a faster disease progression, while decreases in pTau-T181:tau ratio correlate with a slower disease progression. Taken together, alterations in total tau and pTau-T181 levels at the synaptic level as well as in ALS CSF suggest that increases in tau may be associated with a worse prognosis, while increases in pTau-T181 may be associated with a better prognosis. Therefore, CSF tau levels and, more importantly, pTau-T181:tau ratio may serve as a potential biomarker for ALS.

Collectively, our results in a large cohort of human post-mortem mCTX suggest that C-terminus tau is hyperphosphorylated and mis-localized to the nucleus and synapses, though there is no overall increase in tau phosphorylation in ALS. Furthermore, the identification of specific variants in *MAPT* in ALS patients suggest that those mutations may act as disease modifiers and alter the onset and duration of ALS. Lastly, our data provide additional support for the use of pTau-T181:tau ratio as a potential biomarker for ALS.

## Data Availability

The data that support the findings in this study can be requested from the corresponding author and made available pending review of reasonable requests for academic use

## Acknowledgements

T.P. was supported by an award from the Judith and Jean Pape Adams Charitable Foundation and Byrne Family Endowed Fellowship in ALS Research. SD was supported by the Alzheimer’s association (2018-AARF-591935) and the Jack Satter Foundation. S.M.K.F. was supported by the ALS Canada Tim E. Noël Postdoctoral Fellowship. The authors would like to thank the patients and their families for sample donations.

## Author Contributions

T.P. contributed to the study design, data collection, data analysis, and drafting of the manuscript. A.C.A., S.D., S.M.K.F., J.C., B.A.T., P.K., A.N., E.A.B., S.E.K., P.M.D., A.G. contributed to the data collection, data analysis, and editing of the manuscript. T.G.I., B.T.H., S.E.A., T.S.J., M.E.C., J.D.B. contributed to the study design, and editing of the manuscript. GSV contributed to the study design, data analysis, drafting of the manuscript.

---

**Supplementary Table 1.** Tau correlation of pre-post changes.

|  | n | Med_months | ALSFRS-R | tau | p value |
| --- | --- | --- | --- | --- | --- |
| Mean (SD) for change per month from baseline | 18 | 10.7 | -0.32 (0.26) | 0.37 (3.63) | 0.663 |

**Supplementary Table 2.**
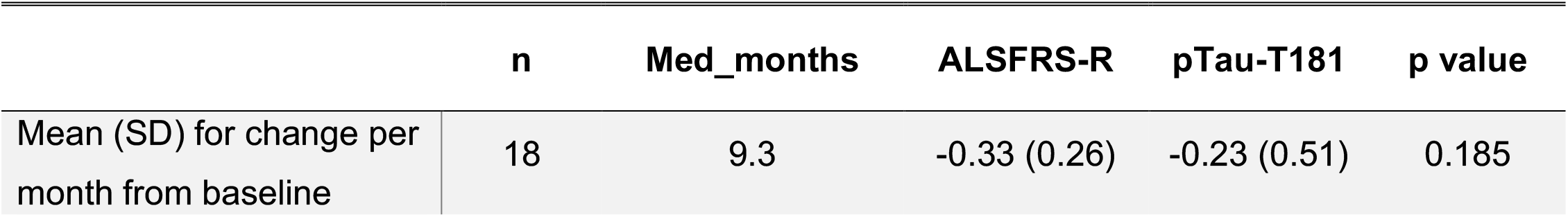
pTau-T181 correlation of pre-post changes.

|  | n | Med_months | ALSFRS-R | pTau-T181 | p value |
| --- | --- | --- | --- | --- | --- |
| Mean (SD) for change per month from baseline | 18 | 9.3 | -0.33 (0.26) | -0.23 (0.51) | 0.185 |

**Supplementary Table 3.** pTau-T181:tau ratio correlation of pre-post changes.

|  | n | Med_months | ALSFRS-R | pTau-T181:tau ratio | p value |
| --- | --- | --- | --- | --- | --- |
| Mean (SD) for change per month from baseline | 18 | 9.3 | -0.32 (0.26) | -0.00 (0.01) | 0.327 |

**Supplementary Figure 1.**
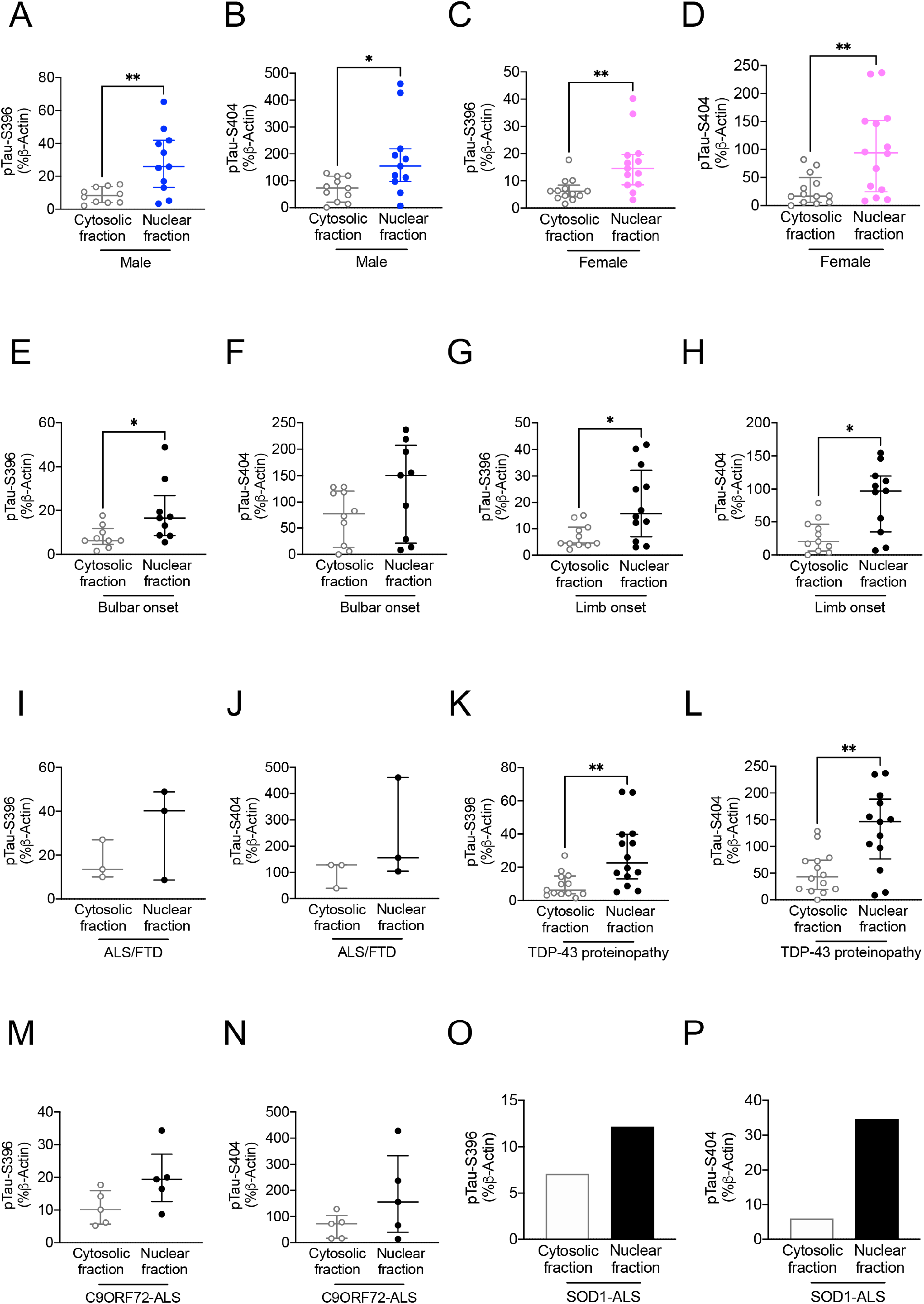
Increases in pTau-S396 and pTau-S404 in ALS nuclear fractions are independent of sex, region of onset, and genotype. (**A**) pTau-S396 levels were significantly increased in the nuclear fraction of male ALS patients (n = 1) (Mann Whitney U = 18, p = 0.0079). (**B**) Nuclear pTau-S404 levels were significantly increased in male ALS patients (n = 11) (Mann Whitney U = 24, p = 0.0158). (**C**) Nuclear pTau-S396 levels were significantly increased in female ALS patients (n = 14) (Mann Whitney U = 29.50, p = 0.0037). (**D**) pTau-S404 levels were significantly increased in the nuclear fraction of female ALS patients (n = 14) (Mann Whitney U = 38, p = 0.0091). (**E**) Nuclear pTau-S396 levels were significantly increased in bulbar onset ALS (n = 10) (Mann Whitney U = 17, p = 0.0379). (**F**) Nuclear pTau-S404 levels were not changed in bulbar onset ALS (n = 10) (Mann Whitney U = 27, p = 0.1564). (**G**) Nuclear pTau-S396 was significantly increased in limb onset ALS (n = 12) (Mann Whitney U = 30, p = 0.0257). (**H**) pTau-S404 levels were significantly increased in the nuclear fraction of limb onset ALS (n = 12) (Mann Whitney U = 22, p = 0.0104). (**I**) Nuclear pTau-S396 levels were not altered in ALS/FTD (n = 3) (Mann Whitney U = 3, p = 0.7000). (**J**) Nuclear pTau-S404 was not altered in ALS/FTD (n = 3) (Mann Whitney U = 2, p = 0.4000). (**K**) Nuclear pTau-S396 levels were significantly increased in ALS mCTX with TDP-43 proteinopathy (n = 14) (Mann Whitney U = 29, p = 0.0019). (**L**) Nuclear pTau-S404 levels were significantly increased in ALS mCTX with TDP-43 proteinopathy (n = 14) (Mann Whitney U = 36, p = 0.0066). (**M**) There was a trend towards a significant increase in nuclear pTau-S396 in *C9ORF72*-ALS (n = 5) (Mann Whitney U = 4, p = 0.0952). (**N**) Nuclear pTau-S404 was not altered in *C9ORF72*-ALS (n = 5) (Mann Whitney U = 8, p = 0.4206). (**O**) Nuclear pTau-S396 levels were increased in a single SOD1-ALS. (**P**) Nuclear pTau-S404 levels were increased in a single SOD1-ALS. The graphs demonstrate the individual integrated density values (IDV) of pTau-S396, and pTau-S404 as % of Β-Actin IDV from western blot experiments performed in nuclear/cytoplasmic fraction in ALS mCTX, with the central line representing the median, and the edges representing the interquartile range. * p < 0.05; ** p < 0.01.

**Supplementary Figure 2.**
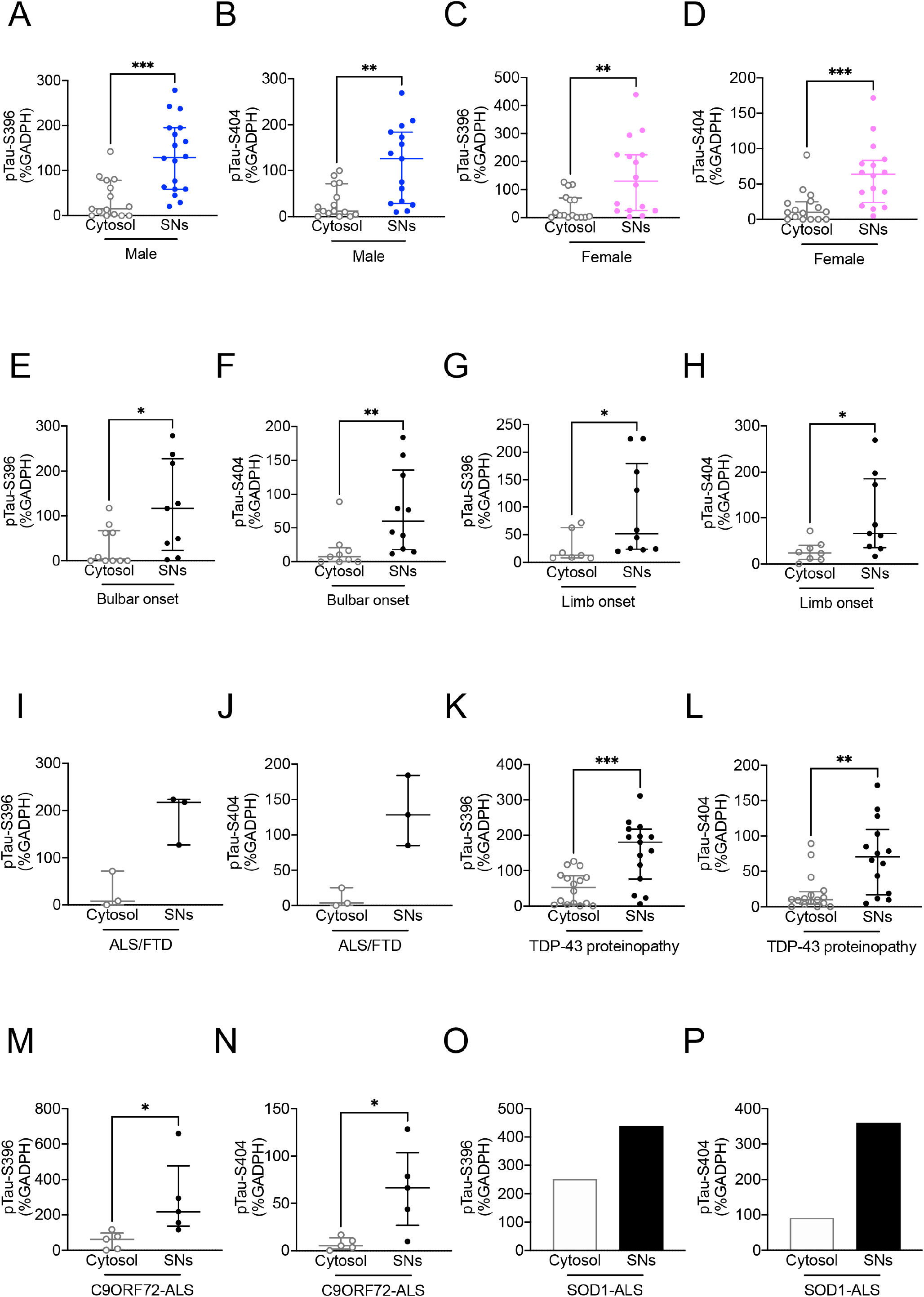
Increases in pTau-S396 and pTau-S404 in ALS SNs are independent of sex, region of onset, and genotype. (**A**) There was a significant shift in pTau-S396 from the cytosolic fraction to the SNs in male ALS patients (n = 20) (Mann Whitney U = 49, p = 0.0004). (**B**) pTau-S404 levels were increased in SNs derived from male ALS patients (n = 20) (Mann Whitney U = 46, p = 0.0014). (**C**) There was a significant increase in pTau-S396 levels in SNs derived from female ALS patients (n = 16) (Mann Whitney U = 42, p = 0.0026). (**D**) pTau-S404 was increased in SNs derived from female ALS patients (n = 16) (Mann Whitney U = 19, p < 0.0001). (**E**) pTau-S396 levels were significantly increased in SNs derived from bulbar onset ALS (n = 13) (Mann Whitney U = 21, p = 0.0023). (**F**) There was a significant shift in pTau-S404 from the cytosol to SNs in bulbar onset ALS (n = 13) (Mann Whitney U = 5.500, p < 0.0001). (**G**) There was a significant shift in pTau-S396 from the cytosol to SNs in limb onset ALS (n = 19) (Mann Whitney U = 58, p = 0.0077). (**H**) pTau-S404 levels were significantly increased in SNs derived from limb onset ALS (n = 19) (Mann Whitney U = 50.50, p = 0.0089). (**I**) There was no significant alteration in pTau-S396 levels in SNs derived from ALS/FTD (n = 3) (Mann Whitney U = 0, p = 0.1000). (**J**) pTau-S404 levels were not altered in SNs derived from ALS/FTD (n = 3) (Mann Whitney U = 0, p = 0.1000). (**K**) There was a significant increase in pTau-S396 levels in SNs derived from ALS with TDP-43 proteinopathy (n = 18) (Mann Whitney U = 46, p = 0.0008). (**L**) pTau-S404 levels were significantly increased in SNs derived from ALS with TDP-43 proteinopathy (n = 18) (Mann Whitney U = 47.50, p = 0.0018). (**M**) There was a significant shift in pTau-S396 from the cytosol to SNs in *C9ORF72*-ALS (n = 5) (Mann Whitney U = 1, p = 0.0159). (**N**) pTau-S404 levels were significantly increased in SNs derived from *C9ORF72*-ALS (n = 5) (Mann Whitney U = 2, p = 0.0317). (**O**) pTau-S396 levels were increased in SNs derived from a single SOD1-ALS. (**P**) pTau-S404 levels were increased in SNs derived from a single SOD1-ALS mCTX. The graphs demonstrate the individual integrated density values (IDV) of pTau-S396, and pTau-S404 as % of GADPH IDV from western blot experiments performed in SNs derived from ALS mCTX, with the central line representing the median, and the edges representing the interquartile range. * p < 0.05; ** p < 0.01, *** p < 0.001, **** p < 0.0001.

**Supplementary Figure 3.**
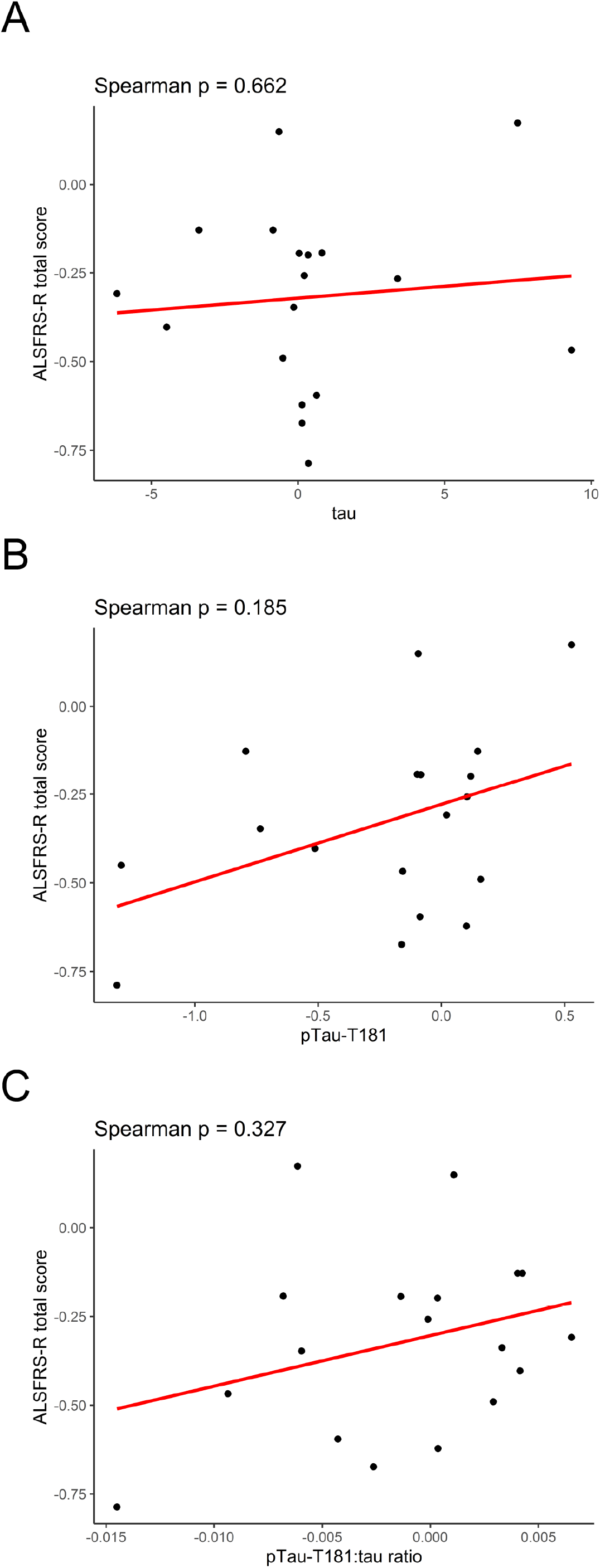
Tau levels and pTau-T181:tau ratio in CSF do not correlate with pre-post changes in ALS. **(A)** Information for tau was available for 18 participants and the median follow up time was 11 months (Spearman correlation, p = 0.662). (**B**) Information for pTau-T181 was available for 18 participants and the median follow up time was 9 months (Spearman correlation, p = 0.185). (**C**) Information for pTau-T181:tau ratio was available for 18 participants and the median follow up time was 9 months (Spearman correlation, p = 0.327). Scatter plots demonstrate the pre-post changes in the given variables. Values were derived from visit 1 to the last visit where all data was available for the given comparisons.

